# Fascioliasis in north-central Vietnam: assessing community knowledge, attitudes, and practices

**DOI:** 10.1101/2024.11.04.24316254

**Authors:** Vinh Hoang Quang, Bruno Levecke, Dung Do Trung, Binh Vu Thi Lam, Le Thuy Dung, Nguyen Duc Thuy, Tran Thi Tuyen, Nguyen Thi Thu Hien, Nguyen Ngoc Ha, Brecht Devleesschauwer, Kathy Goossens, Theodorus de Jong, Linda Paredis, Katja Polman, Steven Callens, Pierre Dorny, Veronique Dermauw

## Abstract

**Background:** Fascioliasis, caused by *Fasciola hepatica* and *Fasciola gigantica*, is a zoonotic disease that significantly impacts public health in agricultural communities, particularly in Vietnam. This study aims to assess the knowledge, attitudes, and practices (KAP) regarding fascioliasis among residents in a rural community in Vietnam.

**Methodology/Principal Findings:** A cross-sectional study was conducted in Dong Thanh commune, north-central Vietnam. A random sample of 621 households was selected, and 1,398 individuals participated in this study. All participants were interviewed for individual KAP to assess KAP regarding fascioliasis. In contrast, household heads were also interviewed about household practices, including life cycle knowledge, health-seeking behavior, water and sanitation practices, livestock and crop management, and dietary habits. Descriptive statistics were used to assess KAP, and generalized linear models were applied to examine the associations between socio-demographic variables and KAP. Awareness of fascioliasis was low, with 85% (1,193/1,398) of respondents reporting no prior knowledge. Detailed understanding of transmission, symptoms, and prevention was limited. Only 9% (124/1,398) of participants could accurately identify the symptoms, while 12% (168/1,398) were knowledgeable about preventive measures. A high percentage of households treated drinking water (99%, 613/619), and consumption of raw vegetables was widespread, with 93% (1,083/1,168) of individuals and 95% of households reporting this practice. Males were less likely to engage in non-risky practices than females (odds ratio: 0.696; 95% confidence interval: 0.591-0.819). Most households (85%, 522/617) sourced plants from their parcels, and 67% (395/588) used animal manure as fertilizer.

**Conclusion/Significance:** The study reveals significant gaps in knowledge, attitudes, and practices related to fascioliasis in Dong Thanh commune. There is a pressing need for targeted educational programs to enhance community awareness and promote safer practices to mitigate the risk of fascioliasis transmission. Future interventions should emphasize gender-specific education and broader community involvement to address these gaps effectively.

**Author Summary:** Fascioliasis, caused by *Fasciola hepatica* and *Fasciola gigantica*, is a zoonotic foodborne disease emerging in Vietnam. This study investigates the knowledge, attitudes, and practices (KAP) related to fascioliasis among residents of a commune in north-central Vietnam, where the disease is highly prevalent in livestock. A cross-sectional survey of 1,398 individuals and 621 households assessed life cycle knowledge, health-seeking behaviors, dietary habits, water and sanitation practices, and management of both livestock and crops. The results show that 85% of respondents were unaware of fascioliasis, with inadequate knowledge about its transmission, symptoms, and prevention. Water treatment is commonly practiced, but consumption of raw vegetables persists. Socio-demographic factors, especially gender, males more likely to engage in risky practices. The findings highlight critical gaps in KAP, emphasizing the need for targeted education to improve awareness and promote safer practices, incorporating gender-specific strategies and community engagement.

## 1. Introduction

Fascioliasis is an ubiquitous parasitic disease caused by the liver flukes *Fasciola hepatica* and *Fasciola gigantica* [1]. It primarily affects ruminants causing economic losses due to significant reductions in milk and meat production [2-5]. Fascioliasis represents a significant public health concern due to its substantial health risks to humans [6, 7]. The World Health Organization (WHO) classifies fascioliasis as a neglected tropical disease, with approximately 50 million individuals infected globally and about 180 million at risk [8]. During the last decades, fascioliasis has become an emerging public health problem in many areas of the world, with an estimated annual loss of 35,000 disability adjusted life years [9]. The disease is acquired through ingesting water or plants contaminated with metacercariae, the parasite’s infective stage, which thrives in environments where human and livestock activities intersect [10].

The northern and central regions of Vietnam are highly endemic for both animal and human fascioliasis [11, 12]. Traditional practices, such as the consumption of raw aquatic plants, facilitate the transmission of the disease to humans [11, 13]. In a comprehensive study by De et al., which analyzed over 53,000 cases of fascioliasis in Vietnam from 1995 to 2019, a notable upward trend in the incidence of the disease was observed. The study documented a rise in the number of cases from a mere two cases reported in 1995 to a peak of 4,026 cases identified in 2019 [12].

In 2020, the General Statistics Office of Vietnam estimated the population of cattle and buffalos at 6.23 and 2.33 million, respectively [14]. In Vietnam, cattle and buffalos are typically managed by small-scale farmers, each maintaining herds of fewer than 30 animals [15], allowing them to freely graze in fields [11]. This practice significantly elevates the risk of infection among these animals, as they are exposed to contaminated environments for the majority of the year [16]. The prevalence of fascioliasis in livestock is high, especially in the Red River Delta area and in Central Vietnam, with infection rates in livestock reaching up to 72% [17].

Despite the clear public health significance of fascioliasis, there is limited information on the community knowledge, attitudes, and practices (KAP) regarding the disease in Vietnam. Understanding these factors is crucial for designing effective intervention strategies. Studies in Iran have shown that awareness and knowledge of fascioliasis are often low in affected communities, leading to behaviors that increase the risk of infection, such as the consumption of raw or improperly treated vegetables and untreated water [18]. Moreover, the utilization of livestock feces as fertilizer and the climatic conditions favoring the presence of suitable snail hosts further perpetuate the persistence and transmission of the disease [1, 19]. Previous research highlighted the critical role of health education and community engagement in controlling zoonotic diseases [17, 20]. Given the rising number of documented fascioliasis cases [12] and the existing gap in comprehensive studies on the disease’s epidemiology and control, this study aims to evaluate the KAP related to fascioliasis among residents of Dong Thanh, a commune in Nghe An province (north-central Vietnam).

## 2. Methods

### 2.1. Ethics statement

This study was approved by the institutional review boards of both the National Institute of Malariology, Parasitology, and Entomology (NIMPE) in Vietnam (Approval No. 02-2022/HDDD) and the Ghent University Hospital/Faculty of Medicine and Health Sciences, Ghent University in Belgium (Approval No. BC-08915). Prior to commencing the fieldwork, the study’s aims and procedures were communicated to provincial and district health officers, as well as the Dong Thanh Commune People’s Committee, which included medical doctors and veterinarians. These local authorities subsequently granted their approval for the study. During the initial visit, detailed information about the study’s objectives and procedures was provided to all participants. Signed informed consent was obtained from each participant aged 18 years and older before they were enrolled in the study. For minors (i.e., participants between 5 and 1 years old), verbal assent was solicited and written informed consent was secured from their parents or guardians.

### 2.2. Study site

This study was conducted in Dong Thanh commune in Yen Thanh district, Nghe An province in north-central Vietnam (**Figure 1**). As of March 2022, this commune has 8,490 registered habitants across 2,036 households. The main occupation of the residents is farming. The selection of this study site was based on (i) historical data on the presence of human fascioliasis, (ii) culinary behaviors that favor disease transmission (e.g. consumption of raw vegetables), (iii) the presence of both cattle and buffalos that have access to crop fields, and (iv) the accessibility of the study site.

**Fig 1.**
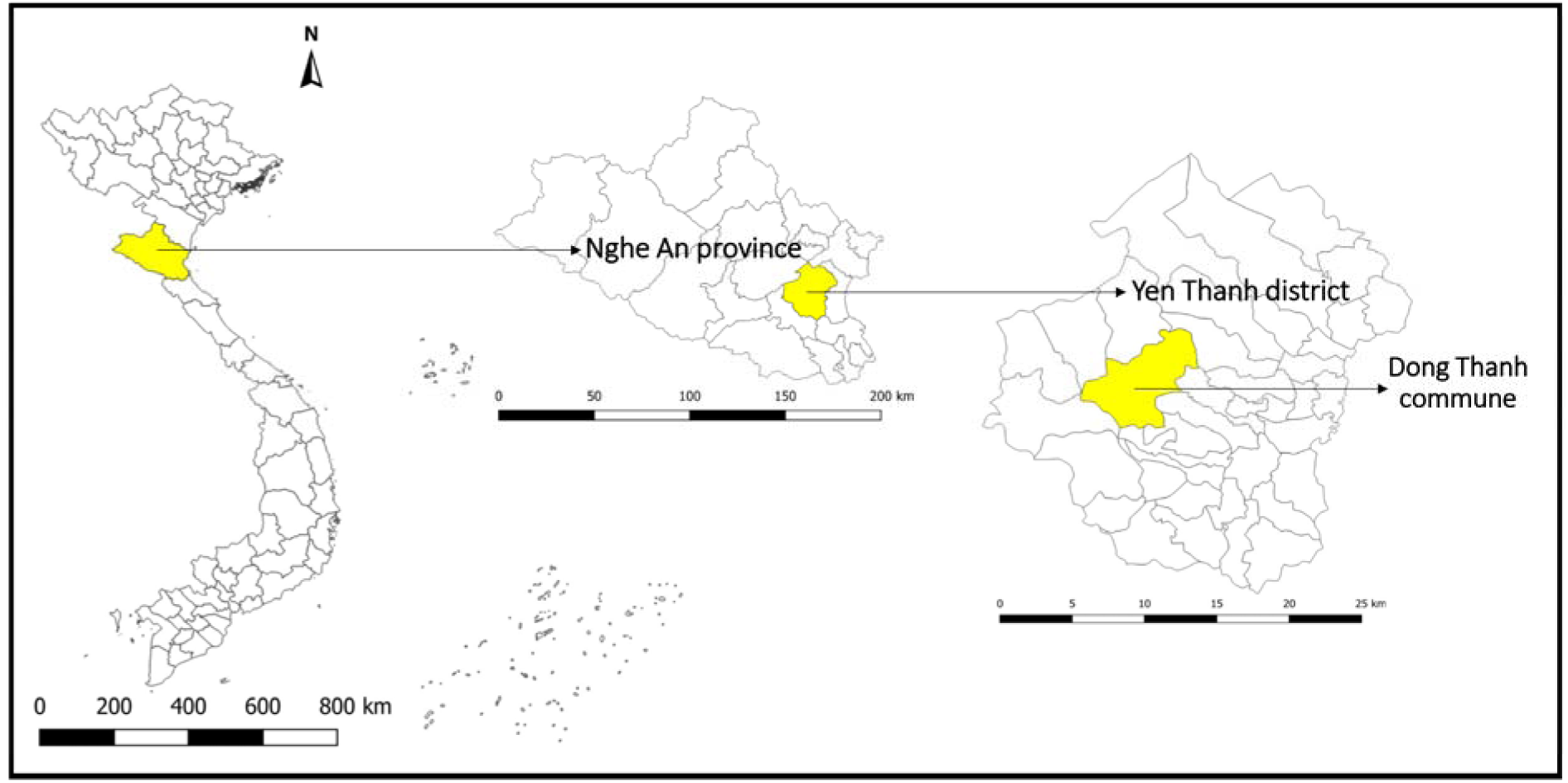
Map of Vietnam showing the locations of Nghe An province, Yen Thanh district, and Dong Thanh commune. The map was created using QGIS Desktop version 3.36.2 software

### 2.3. Study design and data collection

Between April and May 2022, a cross-sectional study was conducted to assess the current KAP related to fascioliasis within the Dong Thanh commune. This study was part of a multi-host study with the primary goal to estimate the prevalence of fascioliasis across all hosts in the life cycle. From a list with the commune’s households provided by the local authorities, households with a minimum of three members were extracted from this list. Subsequently, a random sampling method was applied to this subset to select 621 households. During household visits, if more than three individuals were present, a maximum of three eligible individuals were randomly chosen to participate. In cases when the household comprised three or fewer individuals, all members were invited to participate. After the eligibility of the selected individuals was confirmed, written informed consent was sought for the selected adults, while for children aged 5 to 17, verbal assent combined with written informed consent from their parents or guardians was sought. All selected individuals were invited to participate in the individual questionnaire. The household questionnaire was specifically administered to the household head or the member who possessed the most comprehensive knowledge of household activities. Finally, for all participants, general demographic details were collected including gender, age, occupation, education level, and marital status.

### 2.4. Questionnaires

The individual questionnaire consisted of three comprehensive sections, each designed to capture specific aspects of participants’ KAP regarding fascioliasis. The first section contained questions on general demographic information, including age, gender, education level, and distance from the nearest health clinic. The second section focused on knowledge and awareness of fascioliasis, featuring questions on participants’ familiarity with the disease, its symptoms, modes of transmission, and prevention strategies. The third section explored participants’ attitudes and practices related to fascioliasis, including health-seeking behaviors, dietary habits, and the frequency of consuming raw vegetables. This section also included questions on healthcare access and reasons for not seeking formal medical treatment.

The household questionnaire was structured into four sections (**Info S1**). The first section collected demographic data, including age, gender, occupation, educational attainment, marital status, and participants’ familiarity with fascioliasis. Additionally, the GPS coordinates of each household were recorded in this section. The second section encompassed 11 questions related to household water and sanitation practices. The third section, consisting of 15 questions, focused on livestock and crop management practices. The final section included 8 questions aimed at assessing culinary habits, specifically food preparation methods and the types of plants consumed.

Prior to field implementation, the research team received training on the questionnaire content, conducted pilot interviews among team members, and refined the questions to align with the local language, optimize interview duration, and enhance the clarity and comprehensibility of the phrasing for the local population. Both questionnaires were generated in English and then translated into Vietnamese. The questions were completed by interviewers using a printed version. After the field data collection, the data were entered manually into the RedCap application [21, 22]. Templates of both questionnaires are provided in **Info S1**. All original data are available in **Info S2**.

### 2.5. Statistical data analysis

The data were exported from the RedCap application to an Excel file and consequently analysed by the statistical software R [23]. First, all responses were presented using descriptive statistics. Proportions were shown for categorical variables, whereas mean, median, and range were presented for continuous variables.

The study examined relationships between socio-demographic factors and responses from individual and household questionnaires. For each question, a new binary variable was created: responses indicating non-risky practices or correct knowledge scored 1, while risky practices or incorrect knowledge scored 0 (details in **Info S1**). New variables were created to sum up the 1s and 0s, resulting in a total score for individual practices, household practices, and individual knowledge separately.

Associations between socio-demographic variables (gender, age, education and employment) and individual practice, household practice, and individual knowledge scores were analysed. For individual scores, a generalized linear mixed model with household as a random factor was used. Household practice scores were analysed using a generalized linear model. The full model (i.e., with all the socio-demographic variables included as fixed effects) was simplified by iteratively removing the least significant variable, comparing Akaike’s information criterion values to find the most parsimonious model. The significance level was set at 5%.

## 3. Results

### 3.1. Community KAP of fascioliasis in north-central Vietnam

#### 3.1.1. Socio-demographic characteristics of individual participants

A total of 1,398 individuals were included in the study, with a slight majority of males (55.22%, 770/1,398). The age of the participants ranged from 5 to 92 years, with a mean age of 38 and a median age of 42. Farming was the predominant occupation, accounting for (828/1,397) of respondents. School children and students made up 26.77% (374/1,397) of the sample, while smaller proportions were workers (5.87%, 82/1,397) and government employees (3.44%, 47/1,397). In terms of education, most participants (53.86%, 753/1,398) had completed secondary school. Primary school education was the highest level achieved by 17.38% (243/1,398) of respondents, and 21.75% (304/1,398) had completed high school. A small percentage (5.22%, 73/1,398) had achieved a university degree, and a minority (1.79%, 25/1,398) had not attended school.

#### 3.1.2. Knowledge and attitudes on fascioliasis among individual participants

Knowledge and awareness of fascioliasis among the participants were limited. An important majority (85.34%, 1,193/1,398) had never heard of fascioliasis, while only a small fraction (13.02%, 182/1398) recognized it as a disease affecting humans and livestock. Additionally, 1.57% (22/1,398) of respondents were unsure. Only participants who had heard about the disease or were unsure were asked further knowledge questions. Most of these participants were aware that fascioliasis impacts the liver (77.07%, 158/205). They perceived fascioliasis as somewhat serious (69.95%, 142/203), both at a personal and national level (71.57%, 146/204). Commonly reported symptoms included abdominal pain (44.39%, 91/205) and fever (15.61%, 23/205).

A total of 61.46% (126/205) correctly identified fascioliasis as a parasitic infection, and 60.78% (124/204) understood that it is transmitted through contaminated plants or vegetables. Preventive measures noted included not eating raw water plants (42.44%, 87/205) and cooking vegetables (24.88%, 51/205). There was a strong belief (75.61%, 155/205) that anyone could be infected, and 78.54% (161/205) were aware that specific treatment from health centers could cure the disease. Information sources were primarily television (40.49%, 83/205) and health workers (33.66%, 69/205), while experience with the disease amongst direct contacts, such as neighbors (6.34%; 13/205) and were family members (1.95%; 4/205) were rare. Among 204 respondents, 58.33% (119/204) believed they could contract fascioliasis, 6.37% (13/204) believed they could not, and 35.29% (72/204) were unsure. The majority of the 205 respondents reported that they would primarily feel afraid if given a potential diagnosis (83.90%; 172/205), followed by feeling normal (4.88%; 10/205), surprise (1.95%; 4/205), and sadness or hopelessness (0.49%; 1/205). None of the respondents indicated shame, and 8.78% (18/205) were uncertain about how they would react. More details on the study outcomes can be found in **Info S3**. Because of the low number of participants having responded to more than one knowledge questions (i.e. participants who had heard about fascioliasis), it was impossible to fit a model investigating the association between the socio-demographic characteristics and the individual knowledge scores.

### 3.1.3. Practices on fascioliasis among individual participants

**Table 1** summarizes the healthcare-seeking behavior among 1,398 respondents. A vast majority (95.85%; 1,340/1390) expressed their intention to use health facilities for general health problems, with a small fraction (3.29%; 46/1,398) preferring a pharmacy. A negligible percentage (0.14%; 2/1,398) opted to rest at home, while (3.29%; 46/1,398) of the respondents were uncertain about where they would seek treatment. The frequency of seeking health care was also examined; among 1,340 respondents, (84.55%; 1,133) sought health care at least once a year. Of the 1,392 respondents, (90.88%; 1,265/1392) consumed specific vegetables (shown in pictures), and of the 1,391 respondents, (83.47%; 1,161/1391) indicated they prepare these plants at home. Among 1,168 respondents, (89.81%; 1,049/1,168) consumed raw lettuce, and 56.59% (661/1,168) consumed raw fish mint. Moreover, 60.96% (712/1,168) of these respondents indicated they consume these plants at least once a week. Regarding herbal drink consumption, out of 1,398 respondents, 45.70% (639/1,398) consumed green tea or herbal tea, while 53.29% (745/1,398) did not consume any herbal drinks. Additionally, only 3.22% (45/1,398) of respondents chewed leaves, grass, or other plants found outdoors. When investigating the association between the individual practice scores and the socio-demographic variables, the model containing only gender was best explained the data, with male participants having lower odds for a 1 score, indicating more risky practices, as compared to female participants (odds ratio (OR): 0.696; 95% confidence interval (95%CI): (0.591-0.819)).

**Table 1.**
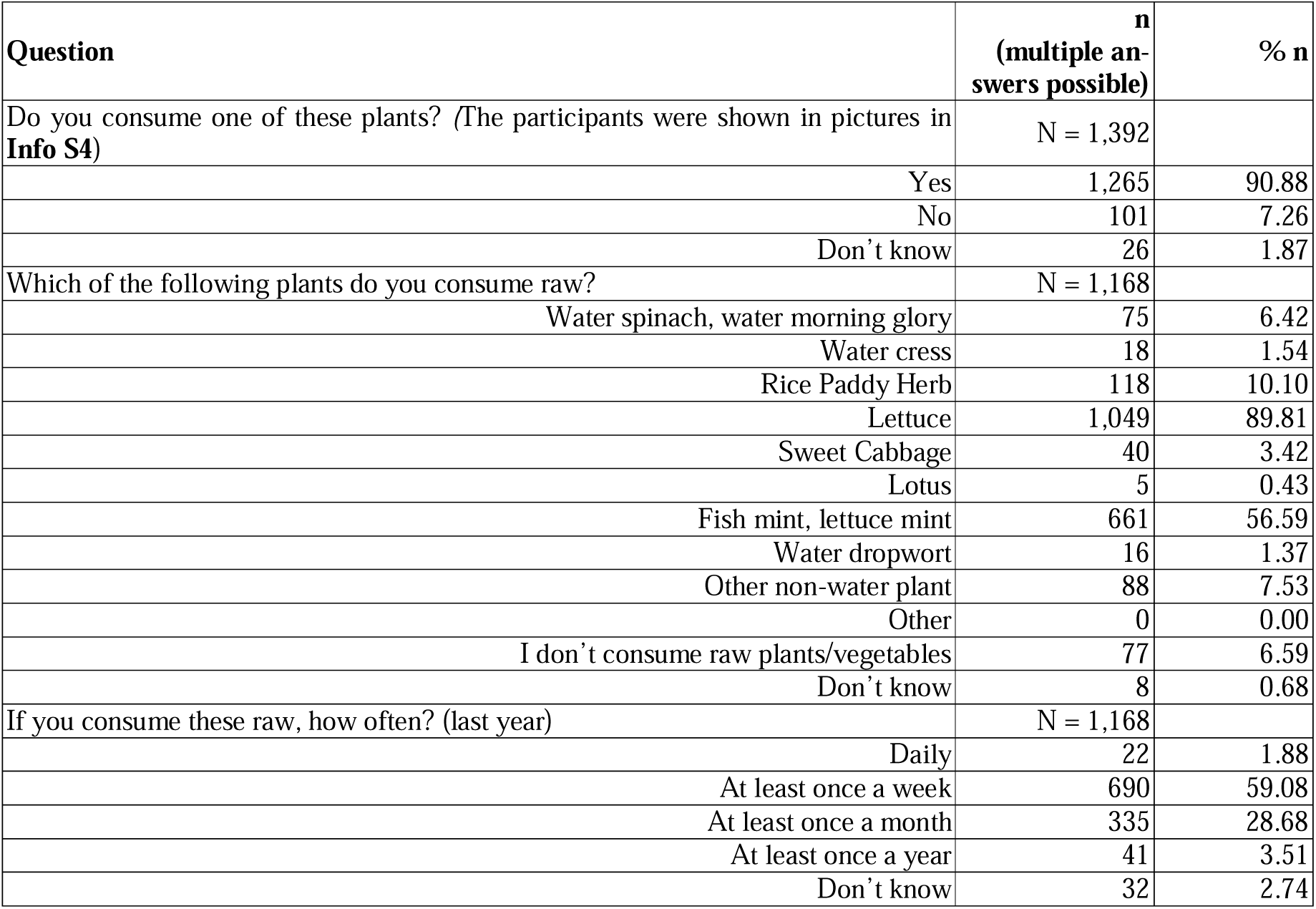
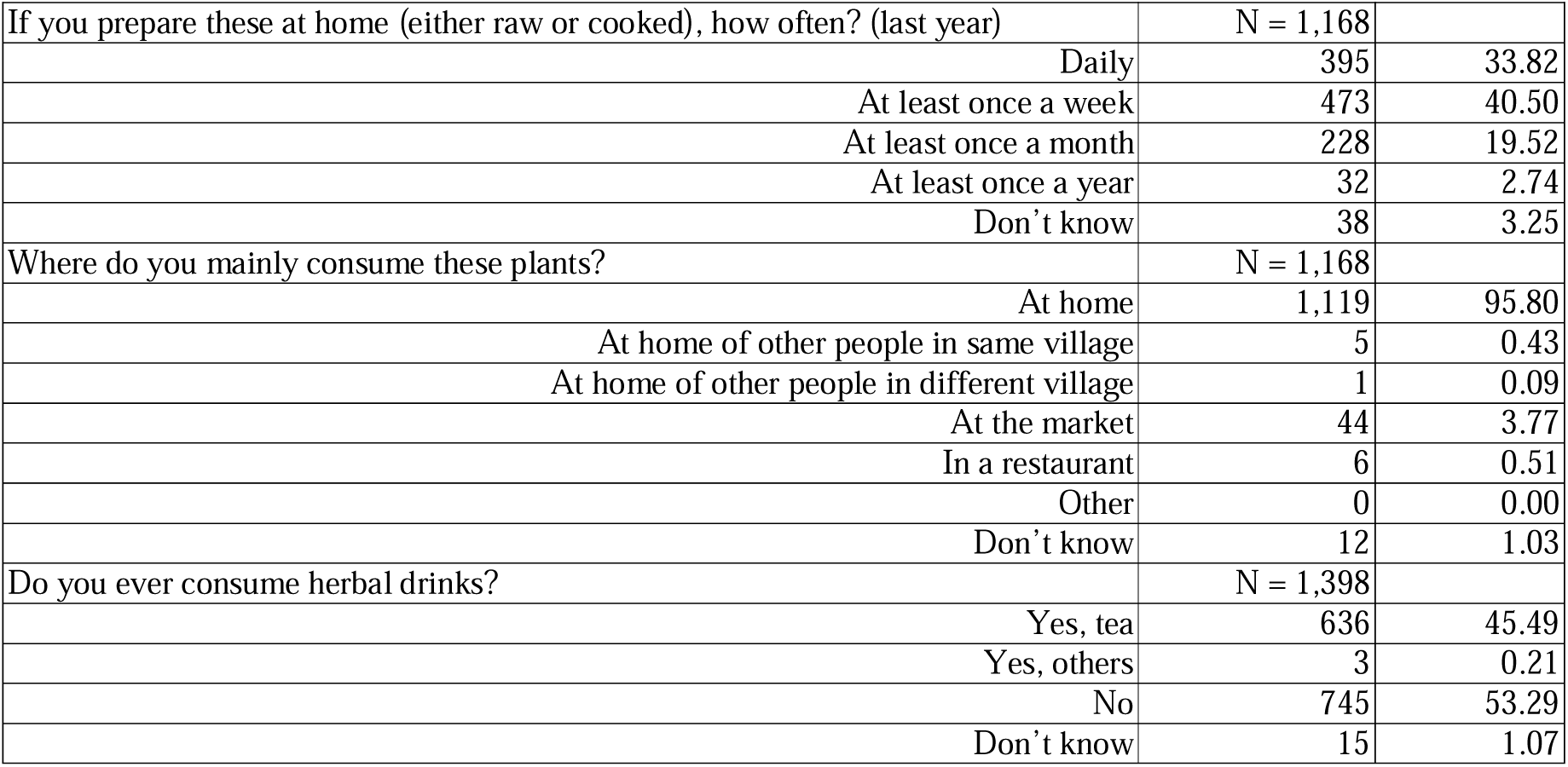
Practices related to fascioliasis among the household participants in Dong Thanh commune.

| Question | n<br>(multiple answers possible) | % n |
| --- | --- | --- |
| Do you consume one of these plants? (The participants were shown in pictures in **Info S4**) | N = 1,392 |  |
| Yes | 1,265 | 90.88 |
| No | 101 | 7.26 |
| Don't know | 26 | 1.87 |
| Which of the following plants do you consume raw? | N = 1,168 |  |
| Water spinach, water morning glory | 75 | 6.42 |
| Water cress | 18 | 1.54 |
| Rice Paddy Herb | 118 | 10.10 |
| Lettuce | 1,049 | 89.81 |
| Sweet Cabbage | 40 | 3.42 |
| Lotus | 5 | 0.43 |
| Fish mint, lettuce mint | 661 | 56.59 |
| Water dropwort | 16 | 1.37 |
| Other non-water plant | 88 | 7.53 |
| Other | 0 | 0.00 |
| I don't consume raw plants/vegetables | 77 | 6.59 |
| Don't know | 8 | 0.68 |
| If you consume these raw, how often? (last year) | N = 1,168 |  |
| Daily | 22 | 1.88 |
| At least once a week | 690 | 59.08 |
| At least once a month | 335 | 28.68 |
| At least once a year | 41 | 3.51 |
| Don't know | 32 | 2.74 |
| If you prepare these at home (either raw or cooked), how often? (last year) | N = 1,168 |  |
| Daily | 395 | 33.82 |
| At least once a week | 473 | 40.50 |
| At least once a month | 228 | 19.52 |
| At least once a year | 32 | 2.74 |
| Don't know | 38 | 3.25 |
| Where do you mainly consume these plants? | N = 1,168 |  |
| At home | 1,119 | 95.80 |
| At home of other people in same village | 5 | 0.43 |
| At home of other people in different village | 1 | 0.09 |
| At the market | 44 | 3.77 |
| In a restaurant | 6 | 0.51 |
| Other | 0 | 0.00 |
| Don't know | 12 | 1.03 |
| Do you ever consume herbal drinks? | N = 1,398 |  |
| Yes, tea | 636 | 45.49 |
| Yes, others | 3 | 0.21 |
| No | 745 | 53.29 |
| Don't know | 15 | 1.07 |

### 3.2. Household practices relating to fasciolosis in north-central, Vietnam

#### 3.2.1. Socio-demographic characteristics of household participants

A total of 621 individuals were interviewed about their household practices. Of the 620 respondents, 64.68% (401/620) were household heads, while 35.32% (219/620) were other household members. The gender distribution among the 621 participants was nearly balanced, with 50.89% (316/621) being male. The age of the participants ranged from 17 to 92 years with a mean of 50 and median age of 52 years. Farming was the primary occupation, representing 84.68% (525/620) of the surveyed individuals. Other occupations included workers (6.94%; 43/620), civil servants or government cadres (3.87%; 24/620), others (4.52%; 28/620), and miscellaneous professions (3.87%). Among the 621 individuals surveyed, education levels varied, ranging from no formal schooling (0.48%; 3/621) to university training (4.67%; 29/621). The largest group (65.70%; 408/621) had completed secondary school, followed by high school graduates (26.09%; 162/621), while 3.06% (19/621) had finished primary education.

#### 3.2.2. Water and sanitation practices in households

Water and sanitation practices are described in **Table 2**. Households primarily relied on tube wells/boreholes (241/621 households, 38.81%) and rainwater collection (239/621 households, 38.49%) for their drinking water, while tube wells/boreholes (339/621 households, 54.59%) and protected dug wells (234/621 households, 37.68%) were water sources used for other purposes in the households. Nearly all households (613/619 households, 98.71%) treated their drinking water, with the majority using filters or water machines (501/615 households, 81.46%). Most households (450/621 households, 72.46%) had access to flush/pour flush toilets, and the wastewater was predominantly managed through septic tanks (454/619 households, 73.34%). Additionally, a vast majority of households (609/617 households, 98.70%) had private toilet facilities.

**Table 2.**
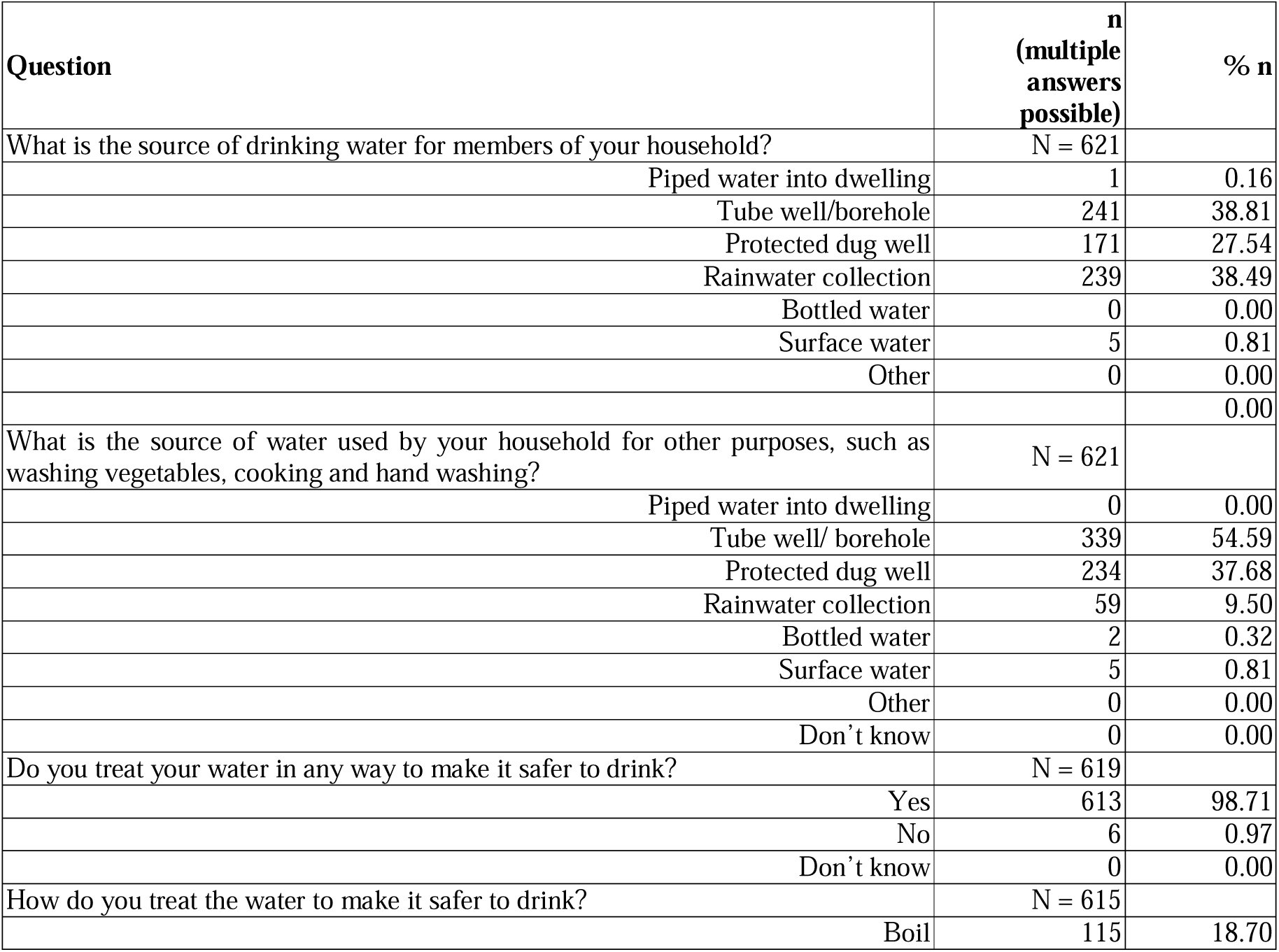
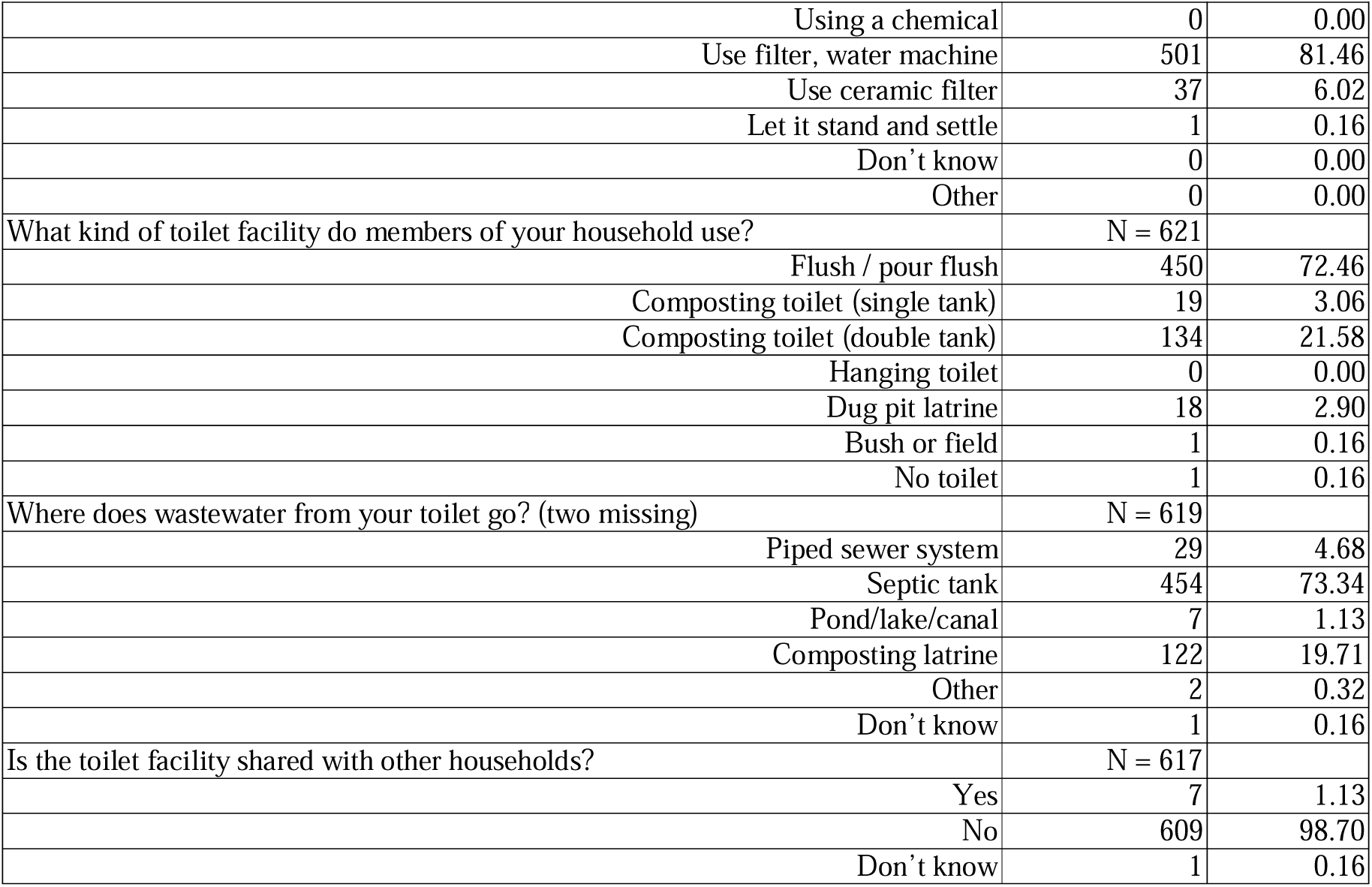
Household water and sanitation practices in Dong Thanh commune.

Related to livestock and crop management (**Table 3**), most households (588/621 households, 94.69%) owned agricultural parcels, with a significant portion (411/585 households, 70.26%) being actively engaged in farming activities. These parcels were predominantly used for crop cultivation, particularly rice, which was the primary crop for most households (565/587 households, 96.25%). Livestock ownership was prevalent, with 567/584 households (96.40%) owning livestock. In terms of livestock interactions with water bodies used for vegetable cultivation, 40/567 respondents (7.05%) reported that their livestock frequently accessed these areas, while 107/567 (18.87%) indicated occasional access. Conversely, 281/567 respondents (49.56%) stated that their livestock never came into contact with such water bodies. Furthermore, 139/567 respondents (24.51%) were uncertain about their livestock’s interaction with these water sources. Among these, 119/567 households (20.99%) owned cattle, and 78/567 households (13.75%) owned buffalo. Manure was commonly utilized as fertilizer (395/588 households, 67.18%), with composting being the preferred treatment method of animal feces (388/395 households, 98.23% of those using manure). Pesticide use was widespread among households (469/587 households, 79.90%), indicating a reliance on chemical inputs for crop management.

**Table 3.**
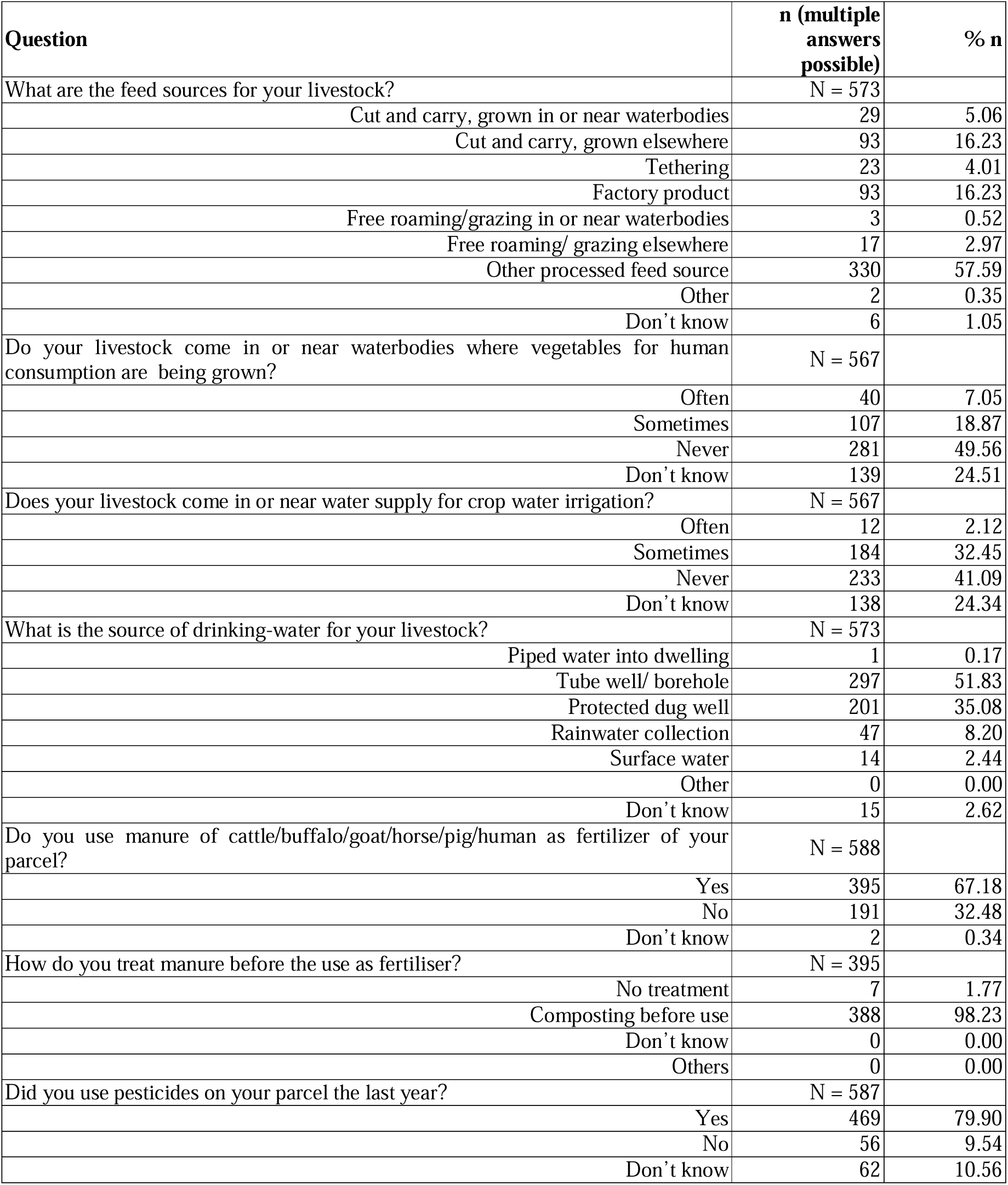
Crop and livestock management practices of households in the Dong Thanh commune.

**Table 4** illustrates the culinary practices among households in the Dong Thanh commune. Most households engaged in the consumption and preparation of various plants. Nearly all households (98.06%; 608/620) consumed these plants, and an equal percentage prepare them at home. Commonly consumed raw plants included lettuce (92.07%; 569/617), fish mint/lettuce mint (64.56%; 399/617), and rice paddy herb (13.43%; 83/617), while the most frequently cooked plants were water spinach/water morning glory (95.30%; 588/617) and sweet cabbage (85.58%; 528/617). Washing practices were prevalent, with 99.67% (616/617) of households washing these plants before use, primarily with water alone (59.48%; 367/617) or with water and salt (40.52%; 250/617). Only (1.3%; 8/617) washed the plants with vinegar and water. Most households (84.60%; 522/617) obtained these plants from their own parcels, while a smaller percentage (17.50%; 108/617) purchased them from local markets (more details can be found in the **Info S3**). None of the investigated socio-demographic variables was found to be significantly associated with the household practice scores.

**Table 4.**
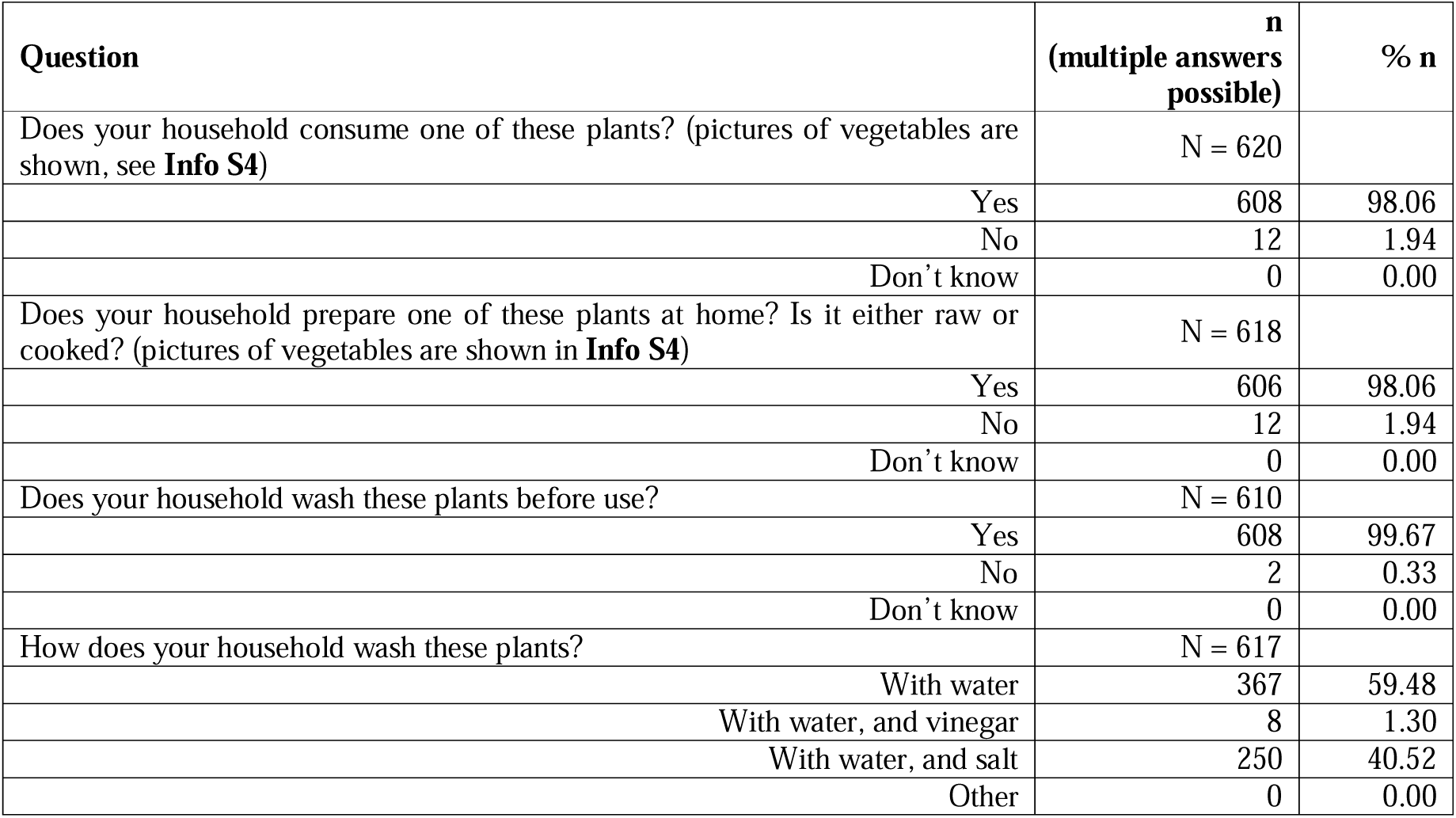
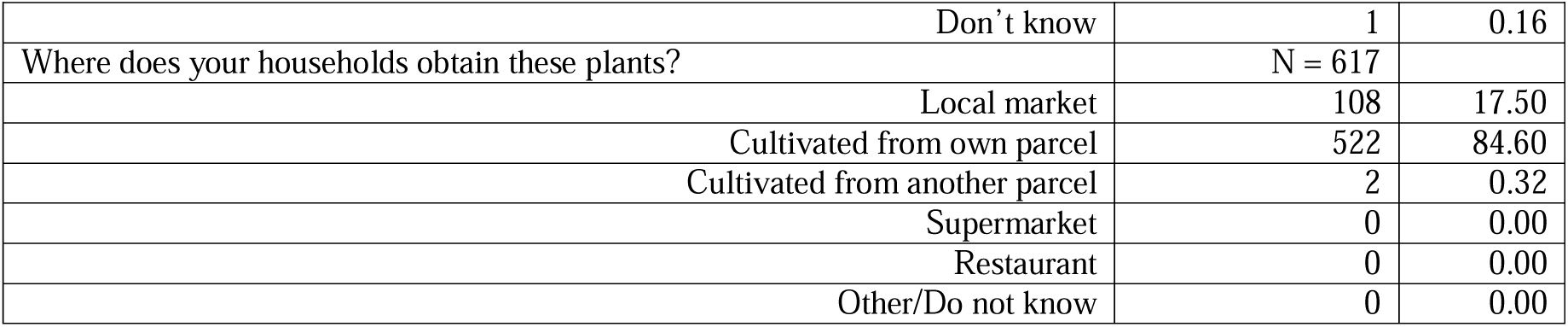
Culinary practices among the households in the Dong Thanh commune.

| Question | n<br>(multiple<br>answers<br>possible) | % n |
| --- | --- | --- |
| Does your household consume one of these plants? (pictures of vegetables are shown, see **Info S4**) | N = 620 |  |
| Yes | 608 | 98.06 |
| No | 12 | 1.94 |
| Don't know | 0 | 0.00 |
| Does your household prepare one of these plants at home? Is it either raw or cooked? (pictures of vegetables are shown in **Info S4**) | N = 618 |  |
| Yes | 606 | 98.06 |
| No | 12 | 1.94 |
| Don't know | 0 | 0.00 |
| Does your household wash these plants before use? | N = 610 |  |
| Yes | 608 | 99.67 |
| No | 2 | 0.33 |
| Don't know | 0 | 0.00 |
| How does your household wash these plants? | N = 617 |  |
| With water | 367 | 59.48 |
| With water, and vinegar | 8 | 1.30 |
| With water, and salt | 250 | 40.52 |
| Other | 0 | 0.00 |
| Don't know | 1 | 0.16 |
| Where does your households obtain these plants? | N = 617 |  |
| Local market | 108 | 17.50 |
| Cultivated from own parcel | 522 | 84.60 |
| Cultivated from another parcel | 2 | 0.32 |
| Supermarket | 0 | 0.00 |
| Restaurant | 0 | 0.00 |
| Other/Do not know | 0 | 0.00 |

## 4. Discussion

Fascioliasis, caused by *F. hepatica* and *F. gigantica*, significantly impacts the health of both humans and animals, mainly as the result of liver damage. This study provides crucial insights into the KAP related to fascioliasis among residents of a rural community in the north-central coast region of Vietnam.

### Lack of knowledge about fascioliasis

A substantial knowledge deficit regarding fascioliasis was revealed in our study, with about 85.34% of participants indicating no prior awareness of the disease. This awareness level is considerably lower compared to findings from other regions. For example, a study in northern Vietnam reported that only 11.50% of participants had never heard of liver fluke [13], consistent with results from Central Vietnam [17, 24, 25]. Another study by Lugong et al., (2024) in the Philippines reported a higher level of awareness, with 81.62% of participants indicating they had heard of fascioliasis [26]. A detailed understanding of fascioliasis’s transmission, symptoms, and prevention among our respondents was notably deficient. This aligns with findings from regions where fascioliasis was prevalent but underreported, indicating a global need for enhanced public health education [1].

### Common practices of consuming raw vegetables and misconceptions about safe handling practices

Our study reveals that the consumption of raw vegetables was frequently practiced within the community. Of the 1,168 respondents, 1,083 (92.72%) indicated consumption of at least one type of raw vegetable. Additionally, 712 participants (61.82%) noted consuming raw vegetables at a minimum frequency of once per week. These findings align with a study by Phi et al. in northern Vietnam, where 91.2% reported consuming raw vegetables [13]. Conversely, Quy et al.’s research found lower rates in central Vietnam, with consumption ranging from 28.20% to 33.80% [17].

In Vietnam, there is a common belief that washing raw vegetables with clean water or soaking them in a saltwater or vinegar solution after washing, makes them safe to eat. Our results showed that 99.84% of households wash vegetables, 59.48% using water, 40.52% saltwater, and 1.30% vinegar water. However, Phuong’s study demonstrated that washing vegetables, whether by rinsing them under running water or scrubbing them by hand, was largely ineffective at removing metacercariae, with less than 2% of the larvae being detached. Additionally, exposure to a 5% acetic acid solution, comparable to commercial vinegar, resulted in 93.60% to 96.00% of metacercariae surviving after less than 20 minutes of treatment. Similarly, soaking vegetables in saltwater for less than 20 minutes failed to effectively kill or detach the metacercariae [27]. These findings are further corroborated by Hassan et al. (2008), who reported that acetic acid solutions below 5% were ineffective at killing *Fasciola* metacercariae, even after 30 minutes of exposure [28].

### Modifying livestock and crop farming practices to reduce fascioliasis transmission

During the KAP study, we concurrently investigated the prevalence of *Fasciola* infection in hosts within the same area. The results revealed a notable discrepancy: animal (buffalo and cattle) infection rates were 51.52% by copro-microscopy and 54.12% by antibody ELISA, while human infection rates were 0% by copro-microscopy and only 0.07% by antibody ELISA [6]. At the same time, this KAP study showed that conditions appear highly conducive to the potential transmission of *Fasciola*. Indeed, this is due to the widespread practice of free-grazing buffalo and cattle in the commune, and people frequently consume raw vegetables, including aquatic plants.

This low infection rate in humans could be explained by several key observations. Firstly, among the raw vegetables consumed, non-aquatic varieties such as lettuce (92.07%) and herbs (64.56%) were the most preferred. Secondly, 84.60% of the vegetables consumed were sourced from household gardens, which included some aquatic plants like fish mint, morning glory, and rice paddy herb. Additionally, households primarily used agricultural land (including submerged land) for rice cultivation (96.25%), with significant portions also allocated to corn (30.66%) and potatoes/sweet potatoes (14.65%). Only a small fraction (3.24%) of the land was dedicated to the cultivation of aquatic vegetables. Furthermore, the study found that out of 395 households using livestock manure in agriculture, 388 households (98.23%) composted the manure before use. In Dong Thanh commune, animal manure is typically collected and piled for 3 to 6 months before being used in agriculture. Composting livestock manure before use helps reduce the risk of *Fasciola* transmission. A study by Moazeni et al. (2010) demonstrated that composting at high temperatures and for a sufficient duration effectively inactivates *Fasciola* eggs and reduces the formation of miracidia [29].

The infection rate of *Fasciola* in the livestock population remains very high, exceeding 50%. This can be attributed to the practice of free grazing of livestock near agricultural fields, which is a significant risk factor for *Fasciola* transmission. Free grazing allows cattle to consume contaminated water and aquatic plants, simultaneously dispersing feces into the agriculture fields. This aligns with previous research by Nguyen et al., which indicated a high risk of infection in livestock due to extensive grazing practices [16].

In general, the practices observed in Dong Thanh commune appear to present significant risks for fascioliasis transmission, particularly due to the high prevalence of infection in livestock, widespread free-grazing, and the consumption of raw vegetables. Combined with the low level of knowledge about the disease, this creates an apparently "dangerous" situation. However, the accompanying prevalence study shows that human infection rates remain very low [30], suggesting that transmission to humans is occurring at a minimal level. Upon closer examination, certain practices, such as the composting of livestock manure, may play a role in reducing the survival of *Fasciola* eggs and limiting transmission risks. Additionally, the practice of growing vegetables in household gardens, with a preference for non-aquatic varieties, may further mitigate potential exposure. Nonetheless, further research is required to gain deeper insights into the specific factors that influence the transmission dynamics and to determine which practices are most effective in reducing the risk of fascioliasis transmission to humans.

### Conclusion

This study reveals significant gaps in KAP related to fascioliasis among residents of Dong Thanh commune, highlighting a substantial lack of awareness. Risky behaviors, such as the frequent consumption of raw vegetables in humans, and free grazing of animals in fields, are prevalent. Although the current prevalence in humans in the area is reported to be low, it is key to stay alert about the potential for transmission, suggesting that certain practices, such as manure composting and the preference for non-aquatic vegetables, may help mitigate transmission. To reduce the risk of fascioliasis transmission, targeted interventions are required. These should include educational programs aimed at raising community awareness about the disease and promoting safer practices in livestock management and vegetable consumption. The role of household gardens in limiting exposure should also be emphasized, alongside improved composting techniques to ensure the inactivation of *Fasciola* eggs. It is essential to incorporate gender-specific strategies and community involvement to enhance the effectiveness of these interventions. Further investigations should elucidate the factors hampering transmission to humans.

## Data Availability

All data produced in the present work are contained in the manuscript

## 5. Acknowledgments

This research was funded by the EmFaVie project, supported by the Flemish Interuniversity Council - University Development Cooperation (VLIR-UOS, grant VN2020SIN317A103 to BL and DDT), and the FasciCoM project, jointly financed by the Research Foundation - Flanders (FWO, grant G0E2921N to BL) and the Vietnamese National Foundation for Science and Technology Development (NAFOSTED, grant FWO.108.2020.01 to DDT). The funders had no role in the study’s design, data collection, analysis, publication decisions, or manuscript preparation. We extend our gratitude to the study participants, the dedicated field team members Nguyen Huu Truong, and Dinh Cong Thinh, and the support from the Nghe An Province Center for Disease Control, Yen Thanh Health District Center, and Dong Thanh commune health stations.

## 7. Supplementary Files

**Info S1** Individual and Household Questionnaires

**Info S2** Original data

**Info S3** Data analysis results

**Info S4** Vegetable pictures

## Notes

### Competing Interest Statement

The authors have declared no competing interest.

### Author Declarations

The institutional review boards of both the National Institute of Malariology, Parasitology, and Entomology (NIMPE) in Vietnam (Approval No. 02-2022/HDDD) and the Ghent University Hospital/Faculty of Medicine and Health Sciences, Ghent University in Belgium (Approval No. BC-08915) gave ethical approval for this work.

