## Supplemental file 1 for "Fascioliasis in north-central Vietnam: assessing community knowledge, attitudes, and practices"

### **Info S1**_**Individual and Household Questionnaires for Human Participants**

### INDIVIDUAL QUESTIONNAIRE

|  | Village | | Household | | | Individual |
| --- | --- | --- | --- | --- | --- | --- |
| ***Individual code*** |  |  |  |  |  |  |

**Part 1: General information**

| 1 | Date (DD/MM/YYYY) |
| --- | --- |
|  | _____/____/_______ |
| 2 | Name |
|  | ______________________________________________________________ |
| 3 | First name(s) |
|  | ______________________________________________________________ |
| 4 | Gender *(circle correct)* |
|  | M / F |
| 5 | Age (in years) |
|  | ______ |
| 6 | Occupation |
|  | School children/student |
|  | Farmer |
|  | Worker |
|  | Cadre, government, civil servant |
|  | Other, specify_________________________________ |
| 7 | Education |
|  | Primary school |
|  | Secondary school |
|  | High school |
|  | University or higher |
|  | Did not go to school |
| 8 | Marital status |
|  | Married |
|  | Single |
|  | Widowed |
|  | Other, specify__________________________________ |
| 9 | How far do you live from the nearest health clinic or hospital? |
|  | 0-10 kilometers |
|  | 11-20 kilometers |
|  | 21-30 kilometers |
|  | More than 30 kilometers |
|  | Don’t know |

**Part 2: Knowledge & awareness**

| Q1 | Have you ever heard about fascioliasis? | Risk score |  |
| --- | --- | --- | --- |
|  | No | 0 | >>Q16 |
|  | Yes, as a human disease | 1 | >>Q2 |
|  | Yes, as a disease in livestock | 1 | >>Q2 |
|  | Yes, as a disease in both humans & livestock | 1 | >>Q2 |
|  | Don’t know | 0 | >>Q2 |
| Q2 | What is fascioliasis? *(multiple choices possible)* | Risk score |  |
|  | A disease affecting skin | 0 | >>Q3 |
|  | A disease affecting liver | 1 | >>Q3 |
|  | A disease affecting bones | 0 | >>Q3 |
|  | A disease affecting heart | 0 | >>Q3 |
|  | Other, specify__________________ | 0 | >>Q3 |
|  | Don’t know | 0 | >>Q3 |

| Q3 | | In your opinion, how serious a disease is fascioliasis? |  |  |
| --- | --- | --- | --- | --- |
|  | | Very serious |  | >>Q4 |
|  | | Somewhat serious |  | >>Q4 |
|  | | Not very serious |  | >>Q4 |
|  | | Don’t know |  | >>Q4 |
| Q4 | In your opinion, how serious a problem is fascioliasis in your country? | | |  |
|  | Very serious | | | >>Q5 |
|  | Somewhat serious | | | >>Q5 |
|  | Not very serious | | | >>Q5 |
|  | Don’t know | | | >>Q5 |

| Q5 | What symptoms does fascioliasis cause? *(multiple choices possible)* | Risk score |  |
| --- | --- | --- | --- |
|  | Headache | 0 | >>Q6 |
|  | Epilepsy | 0 | >>Q6 |
|  | Blurry vision | 0 | >>Q6 |
|  | Backache | 0 | >>Q6 |
|  | Abdominal pain | 1 | >>Q6 |
|  | Fever | 1 | >>Q6 |
|  | Itch | 0 | >>Q6 |
|  | Rash on skin | 0 | >>Q6 |
|  | Jaundice | 1 |  |
|  | Other, specify: _________________________ | N/A | >>Q6 |
|  | Don’t know | 0 | >>Q6 |

| Q6 | What agent is causing fascioliasis? *(multiple choices possible)* | Risk score |  |
| --- | --- | --- | --- |
|  | Bad weather | 0 | >>Q8 |
|  | Lack of nutritious food | 0 | >>Q8 |
|  | You are born with it | 0 | >>Q8 |
|  | A virus | 0 | >>Q7 |
|  | A parasite | 1 | >>Q7 |
|  | A bacteria | 0 | >>Q7 |
|  | Other, specify: ___________________ | 0 | >>Q7 |
|  | Don’t know | 0 | >>Q7 |
| Q7 | How can a person get fascioliasis? *(multiple choices possible)* | Risk score |  |
|  | Through consumption infected organs | 0 | >>Q8 |
|  | Through consumption contaminated plants/vegetables | 1 | >>Q8 |
|  | Through consumption infected meat | 0 | >>Q8 |
|  | Through drinking alcohol | 0 | >>Q8 |
|  | Through consumption of infected fish | 0 | >>Q8 |
|  | Contact with animals | 0 |  |
|  | Other, specify:__________________________ | N/A | >>Q8 |
|  | Don’t know | 0 | >>Q8 |

| Q8 | How can human fascioliasis be prevented? *(multiple choices possible)* | Risk score |  |
| --- | --- | --- | --- |
|  | Isolating infected livestock | 0 | >>Q9 |
|  | Isolating infected humans | 0 | >>Q9 |
|  | Washing water plants | 1 | >>Q9 |
|  | Do not eat raw water plants | 1 | >>Q9 |
|  | Cooking meat or organs | 0 | >>Q9 |
|  | Cooking vegetables \| Nấu chín rau | 1 | >>Q9 |
|  | Treatment of humans | 1 | >>Q9 |
|  | Treatment of animals | 1 | >>Q9 |
|  | It cannot be prevented | 0 | >>Q9 |
|  | Other, specify:___________________ | N/A | >>Q9 |
|  | Don’t know | N/A | >>Q9 |
| Q9 | In your opinion, who can be infected with human fascioliasis? *(multiple choices possible)* | Risk score |  |
|  | Only children | 0 | >>Q10 |
|  | Only poor people | 0 | >>Q10 |
|  | Only homeless people | 0 | >>Q10 |
|  | Only elderly people | 0 | >>Q10 |
|  | Only men | 0 | >>Q10 |
|  | Only women | 0 | >>Q10 |
|  | Anybody | 1 | >>Q10 |
|  | Others, specify:___________________ | 1 | >>Q10 |
|  | Don’t know | 0 | >>Q10 |
| Q10 | Can fascioliasis be cured? *(multiple choices possible)* | Risk score |  |
|  | Yes, with herbal medicine | 0 | >>Q11 |
|  | Yes, home rest without medicine | 0 | >>Q11 |
|  | Yes, specific treatment given by health centre | 1 | >>Q11 |
|  | Yes, using another method, specify:___________________ | N/A | >>Q11 |
|  | It cannot be cured | 0 | >>Q11 |
|  | Don’t know | 0 | >>Q11 |

| Q11 | | Where did you hear about fascioliasis?  *(multiple choices possible)* |  |  |
| --- | --- | --- | --- | --- |
|  | | School |  | >>Q12 |
|  | | Internet |  | >>Q12 |
|  | | Newspapers and magazines |  | >>Q12 |
|  | | Radio |  | >>Q12 |
|  | | TV |  | >>Q12 |
|  | | Billboards/posters |  | >>Q12 |
|  | | Health workers |  | >>Q12 |
|  | | Family, friends, neighbours and colleagues |  | >>Q12 |
|  | | Other, specify:__________________________ |  | >>Q12 |
|  | | Don’t know |  | >>Q12 |
| Q12 | Do you know people who have/had fascioliasis?  *(multiple choices possible)* | | |  |
|  | Yes, I have/had fascioliasis | | | >>Q13 |
|  | Yes, household members | | | >>Q14 |
|  | Yes, relatives (not from household) | | | >>Q14 |
|  | Yes, neighbours | | | >>Q14 |
|  | Yes, friends or colleagues | | | >>Q14 |
|  | Yes, other people, specify:__________________________ | | | >>Q14 |
|  | No | | | >>Q14 |
|  | Don’t know | | | >>Q14 |
| Q13 | | If you have or had fascioliasis, at what point did you go to the health facility? |  |  |
|  | | When treatment on my own didn’t work |  | >>Q14 |
|  | | As soon as I realized that my symptoms might be related with fascioliasis |  | >>Q14 |
|  | | As soon as I was feeling ill |  | >>Q14 |
|  | | I did not go to the health facility |  | >>Q14 |
|  | | Other, specify:__________________________ |  | >>Q14 |
|  | | Don’t know |  | >>Q14 |

**Part 3: Attitudes & practices**

| Q14 | Do you think you could get fascioliasis? |  |  |
| --- | --- | --- | --- |
|  | Yes, because ______________________________________________________________________________________________ |  | >>Q15 |
|  | No, because ______________________________________________________________________________________________ |  | >>Q15 |
|  | Don’t know |  | >>Q15 |

| Q15 | What would be your reaction if you were to found out that you have fascioliasis? *(multiple choices possible)* |  |
| --- | --- | --- |
|  | Normal |  |
|  | Fear | >>Q16 |
|  | Surprise | >>Q16 |
|  | Sadness or hopelessness | >>Q16 |
|  | Shame | >>Q16 |
|  | Other, specify___________________________________ | >>Q16 |
|  | Don’t know | >>Q16 |

| Q16 | Where do you usually go if you are sick, or to treat a general health problem? *(multiple choices possible)* |  |
| --- | --- | --- |
|  | Go to health facility | >>Q18 |
|  | Go to pharmacy | >>Q17 |
|  | Go to traditional healer | >>Q17 |
|  | Pursue other self-treatment options | >>Q17 |
|  | Nowhere, rest at home | >>Q17 |
|  | Other, specify___________________________________ | >>Q17 |
|  | Don’t know | >>Q17 |

| Q17 | | If you would not go to the health facility, what is the reason? *(multiple choices possible)* | |  |
| --- | --- | --- | --- | --- |
|  | | No need | |  |
|  | | Not sure where to go | | >>Q19 |
|  | | Cost | | >>Q19 |
|  | | Difficulties with transportation/distance to clinic | | >>Q19 |
|  | | Do not trust medical workers | | >>Q19 |
|  | | Do not like attitude of medical workers | | >>Q19 |
|  | | Cannot leave work (overlapping work hours with medical facility working hours) | | >>Q19 |
|  | | Do not want to find out something is really wrong | | >>Q19 |
|  | | Other, specify___________________________________ | | >>Q19 |
|  | | Don’t know | | >>Q19 |
| Q18 | | How often do you generally seek health care at a clinic or hospital | |  |
|  | | Twice a year or more | | >>Q19 |
|  | | Once per year | | >>Q19 |
|  | | Less than once a year | | >>Q19 |
|  | | Don’t know | | >>Q19 |
| Q19 | Do you consume one of these plants? *(you will be shown some pictures)* | |  |  |
|  | Yes | |  | >>Q20 |
|  | No | |  | >>Q20 |
|  | Don’t know | |  | >>Q20 |

| Q20 | | Do you prepare one of these plants at home? *(you will be shown some pictures)* | Risk score |  |
| --- | --- | --- | --- | --- |
|  | | Yes | 0 | >>Q21 |
|  | | No | 1 | >>Q25 |
|  | | Don’t know | N/A | >>END |
| Q21 | Which of the following plants do you consume raw? *(you will be shown some pictures)(multiple choices possible)* | | Risk score |  |
|  | Water spinach, water morning glory (Rau muống) | | 0 | >>Q22 |
|  | Water cress (Cải xoong) | | 0 | >>Q22 |
|  | [Rice Paddy Herb](https://web.archive.org/web/20140908201208/http:/www.uni-graz.at/~katzer/engl/Limn_aro.html) (Rau ngổ) | | 0 | >>Q22 |
|  | Salad (Rau xà lách) | | 1 | >>Q22 |
|  | Sweet Cabbage (Rau cải ngọt) | | 1 | >>Q22 |
|  | Lotus (Ngó sen) | | 0 | >>Q22 |
|  | Fish mint, lettuce mint (Rau diếp cá) | | 0 | >>Q22 |
|  | Water dropwort (Rau cần) | | 0 | >>Q22 |
|  | Other non-water plant | | 1 |  |
|  | Others, specify:_____________________________ | | 1 | >>Q22 |
|  | I don’t consume raw plants/vegetables | | 1 | >>Q22 |
|  | Don’t know | | N/A | >>Q22 |

| Q22 | If you consume these raw, how often? (last year) |  |  |
| --- | --- | --- | --- |
|  | Daily |  | >>Q23 |
|  | At least once a week |  | >>Q23 |
|  | At least once a month |  | >>Q23 |
|  | At least once a year |  | >>Q23 |
|  | Don’t know |  | >>Q23 |
| Q23 | If you prepare these at home (either raw or cooked), how often? (last year) |  |  |
|  | Daily |  | >>Q24 |
|  | At least once a week |  | >>Q24 |
|  | At least once a month |  | >>Q24 |
|  | At least once a year |  | >>Q24 |
|  | Don’t know |  | >>Q24 |

| Q24 | Where do you mainly consume these plants? |  |  |
| --- | --- | --- | --- |
|  | At home |  | >>Q25 |
|  | At home of other people in same village |  | >>Q25 |
|  | At home of other people in different village |  | >>Q25 |
|  | At the market |  | >>Q25 |
|  | In a restaurant |  | >>Q25 |
|  | Others, specify:_______________________ |  | >>Q25 |
|  | Don’t know |  | >>Q25 |

| Q25 | Do you ever consume herbal drinks? | Risk score |  |
| --- | --- | --- | --- |
|  | Yes, tea | 0 | >>Q26 |
|  | Yes, others: specify:____________________ | 0 | >>Q26 |
|  | No | 1 | >>Q26 |
|  | Don’t know | N/A | >>Q26 |

| Q26 | Do you ever chew on leaves, grass, or other plants you find outdoors? | Risk score |  |
| --- | --- | --- | --- |
|  | Yes | 0 | >>Q25 |
|  | No | 1 | >>Q25 |
|  | Don’t know | N/A | >>Q25 |

| Q27 | Do you ever consume water chestnut? | Risk score |  |
| --- | --- | --- | --- |
|  | Yes | 1 | >>END |
|  | No | 0 | >>END |
|  | Don’t know | N/A | >>END |

Thank you for your participation!

The Household Questionnaire

Questionnaire – FasciCoM study – Household – Final 15APR22

|  | Village | | Household |
| --- | --- | --- | --- |
| ***HH code*** |  |  |  |

**Part 1 General information**

| 1 | Date (DD/MM/YYYY) |
| --- | --- |
|  | _____/____/_______ |
| 2 | Name |
|  | ______________________________________________________________ |
| 3 | First name(s) |
|  | ______________________________________________________________ |
| 4 | Gender *(circle correct)* |
|  | M / F |
| 5 | Age (in years) |
|  | ______ |
| 6 | Role in household |
|  | Head of household |
|  | Other_______________ |
| 7 | GPS location household |
|  | _____(DDD)° _____(MM)' ____(SS.S)" (latitude)  ____(DDD)° _____(MM)' ____(SS.S)" (longitude) |
| 8 | What is the type of soil of the parcel? *(use handmethod)(only if response to Q29 is: “Cultivated from own parcel or “Cultivated from other parcel (e.g. family and friends)”*) |
|  | Sandy |
|  | Silty |
|  | Clay |
|  | Peaty |
|  | Saline |
|  | Loamy |
|  | Other, specify__________________ |
| 9 | GPS location parcel *(only if response to Q29 is: “Cultivated from own parcel or “Cultivated from other parcel (e.g. family and friends)”*) |
|  | _____(DDD)° _____(MM)' ____(SS.S)" (latitude)  ____(DDD)° _____(MM)' ____(SS.S)" (longitude) |

**Part 2 Water & sanitation practices**

| Q1 | What is the source of drinking-water for members of your household? *(indicate most important one(s))* |  | Risk score |
| --- | --- | --- | --- |
|  | Piped water into dwelling | >>Q2 | 1 |
|  | Tube well/borehole | >>Q2 | 1 |
|  | Protected dug well | >>Q2 | 1 |
|  | Rainwater collection | >>Q2 | 1 |
|  | Bottled water | >>Q2 | 1 |
|  | Surface water (river, dam, lake, pond, stream, canal, irrigation channels) | >>Q1A | 0 |
|  | Other, specify_________________________ | >>Q2 | 0 |
|  | Don’t know | >>Q2 | N/A |

| Q1A | Specify the type of surface water (for drinking)? |  |
| --- | --- | --- |
|  | River | >>Q2B |
|  | Dam | >>Q2B |
|  | Lake | >>Q2B |
|  | Pond, within premises | >>Q2B |
|  | Pond, outside premises | >>Q2B |
|  | Stream | >>Q2B |
|  | Canal | >>Q2B |
|  | Irrigation channels, within premises | >>Q2B |
|  | Irrigation channels, outside premises | >>Q2B |
|  | Mountain water | >>Q2B |
|  | Other, specify:____________________________ | >>Q2B |
|  | Don’t know | >>Q2B |

| Q2 | What is the source of water used by your household for other purposes, such as washing vegetables, cooking and hand washing? *(indicate most important one(s))* |  | Risk score |
| --- | --- | --- | --- |
|  | Piped water into dwelling | >>Q3 | 1 |
|  | Tube well/borehole | >>Q3 | 1 |
|  | Protected dug well | >>Q3 | 1 |
|  | Rainwater collection | >>Q3 | 1 |
|  | Bottled water | >>Q3 | 1 |
|  | Surface water (river, dam, lake, pond, stream, canal, irrigation channels) | >>Q2A | 0 |
|  | Other, specify_____________________ | >>Q3 | N/A |
|  | Don’t know | >>Q3 | N/A |

| Q2A | Specify the type of surface water (for other purposes)? |  |
| --- | --- | --- |
|  | River | >>Q2B |
|  | Dam | >>Q2B |
|  | Lake | >>Q2B |
|  | Pond, within premises | >>Q2B |
|  | Pond, outside premises | >>Q2B |
|  | Stream | >>Q2B |
|  | Canal | >>Q2B |
|  | Irrigation channels, within premises | >>Q2B |
|  | Irrigation channels, outside premises | >>Q2B |
|  | Mountain water | >>Q2B |
|  | Other, specify:____________________________ | >>Q2B |
|  | Don’t know | >>Q2B |

| Q2B | Do livestock have access to this surface water? |  | Risk score |
| --- | --- | --- | --- |
|  | Usually | >>Q3 | 0 |
|  | Often | >>Q3 | 0 |
|  | Sometimes | >>Q3 | 0 |
|  | Never | >>Q3 | 1 |
|  | Don’t know | >>Q3 | N/A |

| Q3 | Do you treat your water in any way to make it safer to drink |  | Risk score |
| --- | --- | --- | --- |
|  | Yes | >>Q4 | 1 |
|  | No | >>Q5 | 0 |
|  | Don’t know | >>Q5 | N/A |

| Q4 | How do you treat the water to make it safer to drink? *(indicate the most important ones)* |  |
| --- | --- | --- |
|  | Boil | >>Q5 |
|  | Using a chemical | >>Q5 |
|  | Use filter, water machine | >>Q5 |
|  | Let it stand and settle | >>Q5 |
|  | Other (specify_____________________) | >>Q5 |
|  | Don’t know | >>Q5 |

| Q5 | What kind of toilet facility do members of your household use? *(indicate most important one(s))* |  | Risk score |
| --- | --- | --- | --- |
|  | Flush/pour flush | >>Q6 | 1 |
|  | Composting toilet (single tank) | >>Q6 | 1 |
|  | Composting toilet (double tanks) | >>Q6 | 1 |
|  | Hanging toilet/hanging latrine/‘Pond toilet’ | >>Q7 | 0 |
|  | Dug pit latrine | >>Q6 | 0 |
|  | Bush or field | >>Q9 | 0 |
|  | Other (specify) | >>Q6 | N/A |
|  | Don’t know | >>Q9 | N/A |
|  | No toilet | >>Q9 | 0 |

**No other specific response**

| Q6 | Where does wastewater from your toilet go? *(indicate most important one(s))* |  | Risk score |
| --- | --- | --- | --- |
|  | Piped sewer system | >>Q7 | 1 |
|  | Septic tank | >>Q7 | 1 |
|  | Pond/lake/canal | >>Q7 | 0 |
|  | Composting latrine | >>Q7 | 1 |
|  | Other, specify: ____________________ | >>Q7 | 0 |
|  | Don’t know | >>Q7 | N/A |

| Q7 | Do you share this facility with other households? |  |
| --- | --- | --- |
|  | Yes | >>Q8 |
|  | No | >>Q9 |
|  | Don’t know | >>Q9 |

| Q8 | How many households use this toilet facility? |  |
| --- | --- | --- |
|  | Other households share this toilet, number: __________________ | >>Q9 |
|  | Any member of the public can use this toilet | >>Q9 |
|  | Don’t know | >>Q9 |

**Part 3 Livestock & crop management**

| Q9 | Do you or any of your household currently own any agriculture parcels exclusively or jointly with someone else? |  |
| --- | --- | --- |
|  | Yes | >>Q10 |
|  | No | >>Q9A |
|  | Don’t know | >>Q9A |

| Q9A | Do you or any of your household work on agriculture parcels? |  |
| --- | --- | --- |
|  | Yes | >>Q10 |
|  | No | >>Q22 |
|  | Don’t know | >>Q22 |

| Q10 | What is the primary use of this parcel? |  |
| --- | --- | --- |
|  | Livestock | >>Q12 |
|  | Crops | >>Q11 |
|  | Livestock and crops | >>Q11 |

| Q11 | What crops are cultivated on this parcel? *(multiple choices are possible)* |  |
| --- | --- | --- |
|  | Rice | >>Q12 |
|  | Water plants | >>Q12 |
|  | Sweet potato/potato | >>Q12 |
|  | Flower | >>Q12 |
|  | Groundnuts | >>Q12 |
|  | Corn | >>Q12 |
|  | Bean | >>Q12 |
|  | Fruit |  |
|  | Other: _______________________________ | >>Q12 |
|  | Don’t know | >>Q12 |

| Q12 | Do you or any of your household currently own any livestock exclusively or jointly with someone else? |  |
| --- | --- | --- |
|  | Yes | >>Q13 |
|  | No | >>Q20 |
|  | Don’t know | >>Q20 |

| Q13 | Which livestock do you own? *(multiple choices are possible)* |  |
| --- | --- | --- |
|  | Cattle | >>Q14 |
|  | Buffalo | >>Q14 |
|  | Goat | >>Q14 |
|  | Pigs | >>Q14 |
|  | Horse | >>Q14 |
|  | Poultry | >>Q14 |
|  | Other, specify:___________________________ | >>Q14 |
|  | Don’t know | >>Q14 |

| Q14 | How many livestock do all members of your household own? *(multiple entries are possible)* |  |
| --- | --- | --- |
|  | Cattle: _________________________ | >>Q15 |
|  | Buffalo : _________________________ | >>Q15 |
|  | Goat: : _________________________ | >>Q15 |
|  | Pigs: _________________________ | >>Q15 |
|  | Horse: _________________________ | >>Q15 |
|  | Poultry: _________________________ | >>Q15 |
|  | Other: _____________________________ | >>Q15 |

| Q15 | What are the feed sources for your livestock? *(indicate most important one(s))* |  | Risk Score |
| --- | --- | --- | --- |
|  | Cut and carry, grown in or near waterbodies | >>Q16 | 0 |
|  | Cut and carry, grown elsewhere | >>Q16 | 1 |
|  | Tethering | >>Q16 | 1 |
|  | Factory product | >>Q16 | 1 |
|  | Free roaming/Grazing, in or near waterbodies | >>Q16 | 0 |
|  | Free roaming/Grazing, elsewhere | >>Q16 | 1 |
|  | Other processed feed source |  | 1 |
|  | Other, specify: ___________________ | >>Q16 | N/A |
|  | Don’t know | >>Q16 | N/A |

| Q16 | What is the purpose of your livestock? *(indicate most important one(s))* |  |
| --- | --- | --- |
|  | Dairy | >>Q17 |
|  | Meat | >>Q17 |
|  | Skin | >>Q17 |
|  | Draft power | >>Q17 |
|  | Sale |  |
|  | Other, specify:___________________ | >>Q17 |
|  | Don’t know | >>Q17 |

| Q17 | Do your livestock come in or near waterbodies where vegetables for human consumption are being grown? |  | Risk Score |
| --- | --- | --- | --- |
|  | Often | >> Q18 | 0 |
|  | Sometimes | >> Q18 | 0 |
|  | Never | >> Q18 | 1 |
|  | Don’t know | >> Q18 | N/A |

| Q18 | Do your livestock come in or near water supply for crop water irrigation? |  | Risk Score |
| --- | --- | --- | --- |
|  | Often | >> Q19 | 0 |
|  | Sometimes | >> Q19 | 0 |
|  | Never | >> Q19 | 1 |
|  | Don’t know | >> Q19 | N/A |

| Q19 | What is the source of drinking-water for your livestock? *(indicate most important one(s))* |  | Risk score |
| --- | --- | --- | --- |
|  | Piped water into dwelling | >>Q20 | 1 |
|  | Tube well/borehole | >>Q20 | 1 |
|  | Protected dug well | >>Q20 | 1 |
|  | Rainwater collection | >>Q20 | 1 |
|  | Surface water (river, dam, lake, pond, stream, canal, irrigation channels) | >>Q20 | 0 |
|  | Other, specify___________________________ | >>Q20 | N/A |
|  | Don’t know | >>Q20 | N/A |

| Q20 | Do you use manure of cattle/buffalo/goat/horse/pig/human as a fertilizer of your parcel? |  | Risk Score |
| --- | --- | --- | --- |
|  | Yes | >>Q20A | 0 |
|  | No | >>Q21 | 1 |
|  | Don’t know | >>Q21 | N/A |

| Q20A | How do you treat manure before use as fertilizer? |  | Risk score |
| --- | --- | --- | --- |
|  | No treatment (use fresh manure) | >>Q21 | 0 |
|  | Composting before use | >>Q21 | 1 |
|  | Others, specify:___________________________ | >>Q21 | N/A |
|  | Don’t know | >>Q21 | N/A |

**No other specific response**

| Q21 | Did you use pesticides on your parcel the last year? |  |
| --- | --- | --- |
|  | Yes, specify type and frequency used the last year: _____________________ | >>Q22 |
|  | No | >>Q22 |
|  | Don’t know_____________________ | >>Q22 |

**Part 4 Culinary practices**

| Q22 | Who is mainly responsible for cooking in this household? |  |
| --- | --- | --- |
|  | Grandmother/father | >>Q23 |
|  | Mother | >>Q23 |
|  | Father | >>Q23 |
|  | Children | >>Q23 |
|  | Others, specify:______________________________ | >>Q23 |
|  | Don’t know | >>Q23 |

| Q23 | Does this household consume one of these plants? *(you will be shown some pictures)* |  | Risk score |
| --- | --- | --- | --- |
|  | Yes | >>Q24 | 0 |
|  | No | >>Q24 | 1 |
|  | Don’t know | >>Q24 | N/A |

| Q24 | Does this household prepare one of these plants at home (either raw or cooked)? *(you will be shown some pictures)* |  | Risk Score |
| --- | --- | --- | --- |
|  | Yes | >>Q25 | 0 |
|  | No | >>END | 1 |
|  | Don’t know | >>END | N/A |

| Q25 | Which of the following plants does your household consume raw? *(you will be shown some pictures)(multiple choices possible)* |  |
| --- | --- | --- |
|  | Water spinach, water morning glory (Rau muống) | >>Q26 |
|  | Water cress (Cải xoong) | >>Q26 |
|  | [Rice Paddy Herb](https://web.archive.org/web/20140908201208/http:/www.uni-graz.at/~katzer/engl/Limn_aro.html) (Rau ngổ) | >>Q26 |
|  | Salad (Rau xà lách) | >>Q26 |
|  | Sweet Cabbage (Rau cải ngọt) | >>Q26 |
|  | Lotus (Ngó sen) | >>Q26 |
|  | Fish mint, lettuce mint (Rau diếp cá) | >>Q26 |
|  | Water dropwort (Rau cần) | >>Q26 |
|  | Other non-waterplants |  |
|  | Others, specify:_____________________________ | >>Q26 |
|  | My household does not consume raw plants/vegetables | >>Q26 |
|  | Don’t know | >>Q26 |

| Q26 | Which of the following plants does your household consume cooked? *(you will be shown some pictures)(multiple choices possible)* |  |
| --- | --- | --- |
|  | Water spinach, water morning glory (Rau muống) | >>Q27 |
|  | Water cress (Cải xoong) | >>Q27 |
|  | [Rice Paddy Herb](https://web.archive.org/web/20140908201208/http:/www.uni-graz.at/~katzer/engl/Limn_aro.html) (Rau ngổ) | >>Q27 |
|  | Salad (Rau xà lách) | >>Q27 |
|  | Sweet Cabbage (Rau cải ngọt) | >>Q27 |
|  | Lotus (Ngó sen) | >>Q27 |
|  | Fish mint, lettuce mint (Rau diếp cá) | >>Q27 |
|  | Water dropwort (Rau cần) | >>Q27 |
|  | Other non-waterplants |  |
|  | Others, specify:_____________________________ | >>Q27 |
|  | My household does not consume cooked plants/vegetables | >>Q27 |
|  | Don’t know | >>Q27 |

| Q27 | Does your household wash these plants before use? |  | Risk score |
| --- | --- | --- | --- |
|  | Yes | >>Q28 | 1 |
|  | No | >>Q29 | 0 |
|  | Don’t know | >>Q28 | N/A |

| Q28 | How does your household wash these plants? *(indicate the most important one(s))* |  | Risk score |
| --- | --- | --- | --- |
|  | With water | >>Q29 | 0 |
|  | With water + vinegar | >>Q29 | 1 |
|  | With water + salt | >>Q29 | 0 |
|  | Other, specify____________________ | >>Q29 | N/A |
|  | Don’t know | >>Q29 | 0 |

| Q29 | Where does your household obtain these plants *(indicate the most important one(s))* |  |
| --- | --- | --- |
|  | Local market (Wet market) | >>END |
|  | Cultivated from own parcel | >>END |
|  | Cultivated from other parcel (e.g. family and friends | >>END |
|  | Super market | >>END |
|  | Restaurant | >>END |
|  | Other, specify____________________ | >>END |
|  | Don’t know | >>END |

Thank you for your participation!
