## Supplemental file 3 for "Fascioliasis in north-central Vietnam: assessing community knowledge, attitudes, and practices"

### Info S3 Data analysis results

### 1.1. Household practices relating to fasciolosis in Northern Vietnam

#### Socio-demographic characteristics of study participants,

A total of 621 individuals, each representing a distinct household, were included in the study. The sample consisted of 316 men (50.9%) and 305 women (49.1%). Participants' ages spanned from 17 to 92 years, with a mean age of 50 and a median age of 52. The majority of participants (408; 65.7%) had attained secondary school education as their highest level of educational achievement. Farming was the predominant occupation, with 525 individuals (84.7%) identifying as farmers.

**Table 1. Socio-demographic characteristics of studied participants from household questionnaire in Dong Thanh commune.**

| **Variable** | **Category** | **Frequency** | **%** |
| --- | --- | --- | --- |
| **Role in the household (N=620)** | Household head | 401 | 64.68 |
|  | Other | 219 | 35.32 |
| **Gender (N=621)** | Male | 316 | 50.89 |
|  | Female | 305 | 49.11 |
| **Occupation (N=620)** | Farmer | 525 | 84.68 |
|  | Worker | 43 | 6.94 |
|  | Civil servant/ government, cadre | 24 | 3.87 |
|  | Other | 28 | 4.52 |
| **Marital status (N=621)** | Married | 597 | 96.14 |
|  | Single | 13 | 2.09 |
|  | widowed | 11 | 1.77 |
| **Education (N=621)** | Did not go to school | 3 | 0.48 |
|  | Primary school | 19 | 3.06 |
|  | Secondary school | 408 | 65.70 |
|  | High school | 162 | 26.09 |
|  | University or higher | 29 | 4.67 |

Water and sanitation practices were described in Table 2, households primarily rely on tube wells/boreholes (241 households, 38.81%) and rainwater collection (239 households, 38.49%) for their drinking water, while tube wells/boreholes (339 households, 54.59%) and protected dug wells (234 households, 37.68%) are the main sources for other household water uses. Nearly all households (613 households, 98.71%) treat their drinking water, with the majority using filters or water machines (501 households, 81.46%). Most households (450 households, 72.46%) have access to flush/pour flush toilets, and the wastewater is predominantly managed through septic tanks (454 households, 73.34%). Additionally, the vast majority of households (609 households, 98.70%) have private toilet facilities, reflecting a strong focus on water treatment and sanitation within the community.

**Table 2. Water and sanitation practices in Dong Thanh commune, Vietnam**

| **Question** | **n (multiple answers possible)** | **% n** |
| --- | --- | --- |
| **What is the source of drinking water for members of your household?** | **N= 621** |  |
| Piped water into dwelling | 1 | 0.16 |
| Tube well/borehole | 241 | 38.81 |
| Protected dug well | 171 | 27.54 |
| Rainwater collection | 239 | 38.49 |
| Bottled water | 0 | 0.00 |
| Surface water | 5 | 0.81 |
| Other | 0 | 0.00 |
|  |  | 0.00 |
| **What is the source of water used by your household for other purposes, such as washing vegetables, cooking and hand washing?** | N= 621 |  |
| Piped water into dwelling | 0 | 0.00 |
| Tube well/ borehole | 339 | 54.59 |
| Protected dug well | 234 | 37.68 |
| Rainwater collection | 59 | 9.50 |
| Bottled water | 2 | 0.32 |
| Surface water | 5 | 0.81 |
| Other | 0 | 0.00 |
| Don’t know | 0 | 0.00 |
| **Do you treat your water in any way to make it safer to drink?** | N= 619 |  |
| Yes | 613 | 98.71 |
| No | 6 | 0.97 |
| Don’t know | 0 | 0.00 |
| **How do you treat the water to make it safer to drink?** | N= 615 |  |
| Boil | 115 | 18.70 |
| Using a chemical | 0 | 0.00 |
| Use filter, water machine | 501 | 81.46 |
| Use ceramic filter | 37 | 6.02 |
| Let it stand and settle | 1 | 0.16 |
| Don’t know | 0 | 0.00 |
| Other | 0 | 0.00 |
| What kind of toilet facility do members of your household use? | N= 621 |  |
| Flush / pour flush | 450 | 72.46 |
| Composting toilet (single tank) | 19 | 3.06 |
| Composting toilet (double tank) | 134 | 21.58 |
| Hanging toilet | 0 | 0.00 |
| Dug pit latrine | 18 | 2.90 |
| Bush or field | 1 | 0.16 |
| No toilet | 1 | 0.16 |
| **Where does wastewater from your toilet go?** | N= 619 |  |
| Piped sewer system | 29 | 4.68 |
| Septic tank | 454 | 73.34 |
| Pond/lake/canal | 7 | 1.13 |
| Composting latrine | 122 | 19.71 |
| Other | 2 | 0.32 |
| Don’t know | 1 | 0.16 |
| **Is the toilet facility shared with other households?** | N= 617 |  |
| Yes | 7 | 1.13 |
| No | 609 | 98.70 |
| Don’t know | 1 | 0.16 |

Livestock and crop management in Table 3: The majority of households (588 households, 94.69%) own agricultural parcels, with a significant portion (411 households, 70.26%) actively engaged in farming activities. These parcels are predominantly used for crop cultivation, particularly rice, which is the primary crop for most households (565 households, 96.25%). Livestock ownership is also prevalent, with 563 households (96.40%) owning livestock and using a variety of feed sources. Primarily, livestock are raised for meat (551 households, 96.16%). Manure is commonly utilized as fertilizer (395 households, 67.18%), with composting being the preferred treatment method (388 households, 98.23% of those using manure). Pesticide use is widespread among households (469 households, 79.90%), indicating a reliance on chemical inputs for crop management. These livestock and agricultural practices demonstrate a robust integration of crop and animal farming, emphasizing the importance of both sectors in the local economy.

**Table 3. Crop and livestock management practices of households in the Dong Thanh commune, Vietnam**

| **Question** | **n (multiple answers possible)** | **% n** |
| --- | --- | --- |
| **Do you or any of your household currently own any agriculture parcels exclusively or joint with someone else?** | N= 621 |  |
| Yes | 588 | 94.69 |
| No | 33 | 5.31 |
| **Do you or anyone from your household work on agriculture parcels**? | N= 31 |  |
| Yes | 4 | 12.90 |
| No | 27 | 87.10 |
| **What is the primary use of this parcel?** | N= 585 |  |
| Livestock | 7 | 1.20 |
| Crops | 411 | 70.26 |
| Livestock and crops | 167 | 28.55 |
| **What crops are cultivated on this parcel?** | N= 587 |  |
| Rice | 565 | 96.25 |
| Water plants | 19 | 3.24 |
| Sweet potato/potato | 86 | 14.65 |
| Flower | 2 | 0.34 |
| Groundnuts | 18 | 3.07 |
| Corn | 180 | 30.66 |
| Bean | 22 | 3.75 |
| Fruit | 17 | 2.90 |
| Other | 20 | 3.41 |
| **Do you or any member of your household currently own any livestock exclusively or jointly with someone else?** | N= 584 |  |
| Yes | 567 | 96.40 |
| No | 21 | 3.60 |
| don’t know | 0 | 0.00 |
| What are the feed sources for your livestock? | N= 573 |  |
| Cut and carry, grown in or near waterbodies | 29 | 5.06 |
| Cut and carry, grown elsewhere | 93 | 16.23 |
| Tethering | 23 | 4.01 |
| Factory product | 93 | 16.23 |
| Free roaming/grazing in or near waterbodies | 3 | 0.52 |
| Free roaming/ grazing elsewhere | 17 | 2.97 |
| Other processed feed source | 330 | 57.59 |
| Other | 2 | 0.35 |
| Don’t know | 6 | 1.05 |
| **What is the purpose of your livestock?** | N= 573 |  |
| Dairy | 0 | 0.00 |
| Meat | 551 | 96.16 |
| Skin | 0 | 0.00 |
| Draft power | 32 | 5.58 |
| Sale | 21 | 3.66 |
| Other | 1 | 0.17 |
| Don’t know | 4 | 0.70 |
| **Do your livestock come in or near waterbodies where vegetables for human consumption are being grown?** | N= 567 |  |
| Often | 40 | 7.05 |
| Sometimes | 107 | 18.87 |
| Never | 281 | 49.56 |
| Don’t know | 139 | 24.51 |
| **Does your livestock come in or near water supply for crop water irrigation?** | N= 567 |  |
| Often | 12 | 2.12 |
| Sometimes | 184 | 32.45 |
| Never | 233 | 41.09 |
| Don’t know | 138 | 24.34 |
| **What is the source of drinking-water for your livestock?** | N= 573 |  |
| Piped water into dwelling | 1 | 0.17 |
| Tube well/ borehole | 297 | 51.83 |
| Protected dug well | 201 | 35.08 |
| Rainwater collection | 47 | 8.20 |
| Surface water | 14 | 2.44 |
| Other | 0 | 0.00 |
| Don’t know | 15 | 2.62 |
| **Do you use manure of cattle/buffalo/goat/horse/pig/human as fertiliser of your parcel?** | N= 588 |  |
| Yes | 395 | 67.18 |
| No | 191 | 32.48 |
| Don’t know | 2 | 0.34 |
| **How do you treat manure before the use as fertiliser?** | N= 395 |  |
| No treatment | 7 | 1.77 |
| Composting before use | 388 | 98.23 |
| Don’t know | 0 | 0.00 |
| Others | 0 | 0.00 |
| **Did you use pesticides on your parcel the last year?** | N= 587 |  |
| Yes | 469 | 79.90 |
| No | 56 | 9.54 |
| Don’t know | 62 | 10.56 |

Culinary practices Table 4: The majority of households engage in the consumption and preparation of various plants. Nearly all households (98.06%) consume these plants, and an equal percentage prepare them at home. Commonly consumed raw plants include salad (92.07%), fish mint/lettuce mint (64.56%), and rice paddy herb (13.43%), while the most frequently cooked plants are water spinach/water morning glory (95.30%) and sweet cabbage (85.58%). Washing practices are prevalent, with 99.67% of households washing these plants before use, primarily with water alone (59.48%) or with water and salt (40.52%). Most households (84.60%) obtain these plants from their own parcels, while a smaller percentage (17.50%) purchase them from local markets.

**Table 4. Culinary practices among the households in the Dong Thanh commune**

| **Question** | **N (multiple answers possible)** | **% n** |
| --- | --- | --- |
| **Does your household consume one of these plants? (pictures of vegetables are shown appendix 3)** | N= 620 |  |
| Yes | 608 | 98.06 |
| No | 12 | 1.94 |
| Don’t know | 0 | 0.00 |
| **Does your household prepare one of these plants at home? Either raw or cooked? (pictures of vegetables are shown in appendix 3)** | N= 618 |  |
| Yes | **N=** 606 | 98.06 |
| No | 12 | 1.94 |
| Don’t know | 0 | 0.00 |
| **Which of the following plants does your household consume raw** | N=617 |  |
| Water spinach, water morning glory | 35 | 5.66 |
| Water cress | 17 | 2.75 |
| [Rice Paddy Herb](https://web.archive.org/web/20140908201208/http:/www.uni-graz.at/~katzer/engl/Limn_aro.html) | 83 | 13.43 |
| Salad | 569 | 92.07 |
| Sweet cabbage | 14 | 2.27 |
| Lotus | 3 | 0.49 |
| Fish mint, lettuce mint | 399 | 64.56 |
| Water dropwort | 5 | 0.81 |
| Other non-waterplants | 36 | 5.83 |
| Other | 0 | 0.00 |
| Don’t know | 0 | 0.00 |
| My household does not consume raw plants/vegetables | 28 | 4.53 |
| **Which of the following plants does your household consume cooked?** | N=617 |  |
| Water spinach, water morning glory | 588 | 95.30 |
| Water cress | 72 | 11.67 |
| [Rice Paddy Herb](https://web.archive.org/web/20140908201208/http:/www.uni-graz.at/~katzer/engl/Limn_aro.html) | 75 | 12.16 |
| Salad | 14 | 2.27 |
| Sweet cabbage | 528 | 85.58 |
| Lotus | 0 | 0.00 |
| Fish mint, lettuce mint | 4 | 0.65 |
| Water dropwort | 176 | 28.53 |
| Other non-waterplants | 44 | 7.13 |
| Other | 0 | 0.00 |
| Don’t know | 0 | 0.00 |
| My household does not consume cooked plants/vegetables | 0 | 0.00 |
| **Does your household wash these plants before use?** | N= 610 |  |
| Yes | 608 | 99.67 |
| No | 2 | 0.33 |
| Don’t know | 0 | 0.00 |
| **How does your household wash these plants?** | N= 617 |  |
| With water | 367 | 59.48 |
| With water + vinegar | 8 | 1.30 |
| With water + salt | 250 | 40.52 |
| Other | 0 | 0.00 |
| Don’t know | 1 | 0.16 |
| Where does your households obtain these plants? | N= 617 |  |
| Local market | 108 | 17.50 |
| Cultivated from own parcel | 522 | 84.60 |
| Cultivated from another parcel | 2 | 0.32 |
| Super market | 0 | 0.00 |
| Restaurant | 0 | 0.00 |
| Other | 0 | 0.00 |
| Don’t know | 0 | 0.00 |

### 1.2. Community knowledge, attitudes, and practices of Fascioliasis in northern Vietnam

A total of 1,398 individuals were included in the study, with a slight majority of males (55.22%) over females (44.78%). Farming was the predominant occupation, accounting for 59.27% of the 1,397 respondents. School children and students made up 26.77% of the sample, while smaller proportions were workers (5.87%) and government employees (3.44%). In terms of education, the majority of participants (53.86%) had completed secondary school. Primary school education was the highest level achieved by 17.38% of respondents, and 21.75% had completed high school. A small percentage (5.22%) had attained university-level education or higher, and a minority (1.79%) had not attended school.

Table 5 shows awareness and knowledge of fascioliasis among participants are limited. A majority (85.40%) have never heard of fascioliasis, while only a small fraction (7.09%) recognize it as a disease affecting both humans and livestock. Understanding of the disease is primarily centered on its impact on the liver (77.07%). Most participants perceive fascioliasis as somewhat serious (69.95%), both personally and as a national concern (71.57%). Commonly identified symptoms include abdominal pain (44.39%) and fever (15.61%). Knowledge about the cause of fascioliasis reveals that 61.46% correctly identify it as a parasitic infection, and 60.78% understand it is transmitted through contaminated plants or vegetables. Preventive measures noted include not eating raw water plants (42.44%) and cooking vegetables (24.88%). There is a strong belief (75.61%) that anyone can be infected, and 78.54% are aware that specific treatment from health centers can cure the disease. Information sources are primarily TV (40.49%) and health workers (33.66%), though personal connections and direct experiences with fascioliasis are rare, with only 6.34% knowing neighbors affected by it.

**Table 5. Participant knowledge & awareness of fascioliasis among participants in the Dong Thanh commune.**

| **Question** | **n (multiple answers possible)** | **% n** |
| --- | --- | --- |
| **Have you ever heard about fascioliasis?** | **N=** 1,397 |  |
| No | 1,193 | 85.40 |
| Yes, as a human disease | 66 | 4.72 |
| Yes, as a disease in livestock | 17 | 1.22 |
| Yes, as a disease in both humans & livestock | 99 | 7.09 |
| Don’t know | 22 | 1.57 |
| **What is fascioliasis?** *(multiple choices possible)* | 205 |  |
| A disease affecting skin | 7 | 3.41 |
| A disease affecting liver | 158 | 77.07 |
| A disease affecting bones | 1 | 0.49 |
| A disease affecting heart | 2 | 0.98 |
| Other | 2 | 0.98 |
| Don’t know | 37 | 18.05 |
| **In your opinion, how serious a disease is fascioliasis?** | **N=** 203 |  |
| Very serious | 41 | 20.20 |
| Somewhat serious | 142 | 69.95 |
| Not very serious | 3 | 1.48 |
| Don’t know | 17 | 8.37 |
| **In your opinion, how serious a problem is fascioliasis in your country?** | 204 |  |
| Very serious | 38 | 18.63 |
| Somewhat serious | 146 | 71.57 |
| Not very serious | 3 | 1.47 |
| Don’t know | 17 | 8.33 |
| **What symptoms does fascioliasis cause?** | **N=** 205 |  |
| Headache | 32 | 15.61 |
| Epilepsy | 5 | 2.44 |
| Blurry vision | 6 | 2.93 |
| Backache | 6 | 2.93 |
| Abdominal pain | 91 | 44.39 |
| Fever | 32 | 15.61 |
| Itch | 24 | 11.71 |
| Rash on skin | 9 | 4.39 |
| Jaundice | 1 | 0.49 |
| Other | 6 | 2.93 |
| Don’t know | 82 | 40.00 |
| **What agent is causing fascioliasis?** | **N=** 205 |  |
| Bad weather | 1 | 0.49 |
| Lack of nutritious food | 0 | 0.00 |
| You are born with it | 0 | 0.00 |
| A virus | 8 | 3.90 |
| A parasite | 126 | 61.46 |
| A bacteria | 6 | 2.93 |
| Other | 1 | 0.49 |
| Don’t know | 64 | 31.22 |
| **How can a person get fascioliasis?** | **N=** 204 |  |
| Through consumption infected organs | 17 | 8.33 |
| Through consumption contaminated plants/vegetables | 124 | 60.78 |
| Through consumption infected meat | 14 | 6.86 |
| Through drinking alcohol | 0 | 0.00 |
| Through consumption of infected fish | 2 | 0.98 |
| Contact with animals | 3 | 1.47 |
| Other | 4 | 1.96 |
| Don’t know | 62 | 30.39 |
| **How can human fascioliasis be prevented?** | **N=** 205 |  |
| Isolating infected livestock | 4 | 1.95 |
| Isolating infected humans | 1 | 0.49 |
| Washing water plants | 24 | 11.71 |
| Do not eat raw water plants | 87 | 42.44 |
| Cooking meat or organs | 37 | 18.05 |
| Cooking vegetables \| Nấu chín rau | 51 | 24.88 |
| Treatment of humans | 4 | 1.95 |
| Treatment of animals | 2 | 0.98 |
| It cannot be prevented | 0 | 0.00 |
| Other | 0 | 0.00 |
| Don’t know | 56 | 27.32 |
| **In your opinion, who can be infected with human fascioliasis?** | **N=** 205 |  |
| Only children | 6 | 2.93 |
| Only poor people | 0 | 0.00 |
| Only homeless people | 1 | 0.49 |
| Only elderly people | 6 | 2.93 |
| Only men | 3 | 1.46 |
| Only women | 0 | 0.00 |
| Anybody | 155 | 75.61 |
| Other | 1 | 0.49 |
| Don’t know | 35 | 17.07 |
| **Can fascioliasis be cured?** | **N=** 205 |  |
| Yes, with herbal medicine | 2 | 0.98 |
| Yes, home rest without medicine | 8 | 3.90 |
| Yes, specific treatment given by health centre | 161 | 78.54 |
| Yes, using another method, specify:___________________ | 0 | 0.00 |
| It cannot be cured | 5 | 2.44 |
| Don’t know | 30 | 14.63 |
| **Where did you hear about fascioliasis?** | **N=** 205 |  |
| *(multiple choices possible)* |  | 0.00 |
| School | 11 | 5.37 |
| Internet | 4 | 1.95 |
| Newspapers and magazines | 50 | 24.39 |
| Radio | 53 | 25.85 |
| TV | 83 | 40.49 |
| Billboards/posters | 1 | 0.49 |
| Health workers | 69 | 33.66 |
| Family, friends, neighbours and colleagues | 27 | 13.17 |
| Other | 2 | 0.98 |
| Don’t know | 16 | 7.80 |
| **Do you know people who have/had fascioliasis?** | **N=** 205 |  |
| Yes, I have/had fascioliasis | 4 | 1.95 |
| Yes, household members | 4 | 1.95 |
| Yes, relatives (not from household) | 6 | 2.93 |
| Yes, neighbours | 13 | 6.34 |
| Yes, friends or colleagues | 1 | 0.49 |
| Yes, other people | 0 | 0.00 |
| No | 90 | 43.90 |
| Don’t know | 87 | 42.44 |
| **If you have or had fascioliasis, at what point did you go to the health facility?** | **N=** 4 |  |
| When treatment on my own didn’t work | 0 | 0.00 |
| As soon as I realized that my symptoms might be related with fascioliasis | 0 | 0.00 |
| As soon as I was feeling ill | 3 | 1.46 |
| I did not go to the health facility | 1 | 0.49 |
| Other | 0 | 0.00 |
| Don’t know | 0 | 0.00 |

The data in Table 6 indicated that health-seeking behavior was assessed among 1,398 respondents. A significant majority, 95.85%, reported that they would go to a health facility for general health problems, while 3.29% preferred to visit pharmacies. The frequency of seeking health care was also examined; among 1,340 respondents, 84.55% sought health care at least once a year. The consumption and preparation of water plants were key areas of interest. Of the 1,392 respondents, 90.88% consumed specific vegetables (shown in pictures), and 83.47% of 1,391 respondents reported preparing these plants at home. Among 1,168 respondents, 89.81% consumed salad raw, and 56.59% consumed fish mint raw. Furthermore, 59.08% of these respondents reported consuming these plants at least once a week. Regarding herbal drink consumption, out of 1,398 respondents, 45.70% consumed green tea or herbal tea, while 53.29% did not consume any herbal drinks. Additionally, only 3.22% of respondents reported chewing leaves, grass, or other plants found outdoors. Water chestnut consumption was notably low, with only 0.22% of the 1,392 respondents reporting consumption, and 22.41% were unsure.

**Table 6. Attitudes and practices related to fascioliasis among the participants in Dong Thanh commune**

| **Question** | **n (multiple answers possible)** | **% n** |
| --- | --- | --- |
| Do you think you could get fascioliasis? | **N=** 204 |  |
| Yes, because | 119 | 58.33 |
| No, because | 13 | 6.37 |
| Don’t know | 72 | 35.29 |
| **What would be your reaction if you were to find out that you have fascioliasis** | 205 |  |
| Normal | 10 | 4.88 |
| Fear | 172 | 83.90 |
| Surprise | 4 | 1.95 |
| Sadness or hopelessness | 1 | 0.49 |
| Shame | 0 | 0.00 |
| Other | 1 | 0.49 |
| Don’t know | 18 | 8.78 |
| **Where do you usually go if you are sick, or to treat a general health problem?** | **N=** 1,398 |  |
| Go to health facility | 1,340 | 95.85 |
| Go to pharmacy | 46 | 3.29 |
| Go to traditional healer | 0 | 0.00 |
| Pursue other self-treatment options | 0 | 0.00 |
| Nowhere, rest at home | 2 | 0.14 |
| Other | 0 | 0.00 |
| Don’t know | 46 | 3.29 |
| **If you would not go to the health facility, what is the reason?** | 58 |  |
| No need | 2 | 3.45 |
| Not sure where to go | 2 | 3.45 |
| Cost | 1 | 1.72 |
| Difficulties with transportation/distance to clinic | 3 | 5.17 |
| Do not trust medical workers | 0 | 0.00 |
| Do not like attitude of medical workers | 0 | 0.00 |
| Cannot leave work (overlapping work hours with medical facility working hours) | 2 | 3.45 |
| Do not want to find out something is really wrong | 0 | 0.00 |
| Other | 0 | 0.00 |
| Don’t know | 37 | 63.79 |
| **How often do you generally seek health care at a clinic or hospital** | **N=** 1,340 |  |
| Twice a year or more | 612 | 45.67 |
| Once per year | 521 | 38.88 |
| Less than once a year | 183 | 13.66 |
| Don’t know | 24 | 1.79 |
| **Do you consume one of these plants?** *(you will be shown some pictures)* | 1,392 |  |
| Yes | 1,265 | 90.88 |
| No | 101 | 7.26 |
| Don’t know | 26 | 1.87 |
| Do you prepare one of these plants at home? | **N=** 1,391 |  |
| Yes | 1,161 | 83.47 |
| No | 204 | 14.67 |
| Don’t know | 26 | 1.87 |
| **Which of the following plants do you consume raw?** | **N=** 1,168 |  |
| Water spinach, water morning glory | 75 | 6.42 |
| Water cress | 18 | 1.54 |
| [Rice Paddy Herb](https://web.archive.org/web/20140908201208/http:/www.uni-graz.at/~katzer/engl/Limn_aro.html) | 118 | 10.10 |
| Salad | 1,049 | 89.81 |
| Sweet Cabbage | 40 | 3.42 |
| Lotus | 5 | 0.43 |
| Fish mint, lettuce mint | 661 | 56.59 |
| Water dropwort | 16 | 1.37 |
| Other non-water plant | 88 | 7.53 |
| Other | 0 | 0.00 |
| I don’t consume raw plants/vegetables | 77 | 6.59 |
| Don’t know | 8 | 0.68 |
| **If you consume these raw, how often? (last year)** | **N=** 1,168 | 100.00 |
| Daily | 22 | 1.88 |
| At least once a week | 690 | 59.08 |
| At least once a month | 335 | 28.68 |
| At least once a year | 41 | 3.51 |
| Don’t know | 32 | 2.74 |
| **If you prepare these at home (either raw or cooked), how often? (last year)** | **N=** 1,168 | 100.00 |
| Daily | 395 | 33.82 |
| At least once a week | 473 | 40.50 |
| At least once a month | 228 | 19.52 |
| At least once a year | 32 | 2.74 |
| Don’t know | 38 | 3.25 |
| **Where do you mainly consume these plants?** | **N=** 1,168 | 100.00 |
| At home | 1,119 | 95.80 |
| At home of other people in same village | 5 | 0.43 |
| At home of other people in different village | 1 | 0.09 |
| At the market | 44 | 3.77 |
| In a restaurant | 6 | 0.51 |
| Other | 0 | 0.00 |
| Don’t know | 12 | 1.03 |
| **Do you ever consume herbal drinks?** | 1,398 | 100.00 |
| Yes, tea | 636 | 45.49 |
| Yes, others | 3 | 0.21 |
| No | 745 | 53.29 |
| Don’t know | 15 | 1.07 |
| **Do you ever chew on leaves, grass, or other plants you find outdoors?** | 1,398 | 100.00 |
| Yes | 45 | 3.22 |
| No | 1,336 | 95.57 |
| Don’t know | 17 | 1.22 |
| **Do you ever consume water chestnut?** | **N=** 1,392 | 100.00 |
| Yes | 3 | 0.22 |
| No | 1,077 | 77.37 |
| Don’t know | 312 | 22.41 |
