## Supplemental file 4 for "Fascioliasis in north-central Vietnam: assessing community knowledge, attitudes, and practices"

### Water plants

| | | |
|---|---|---|
| 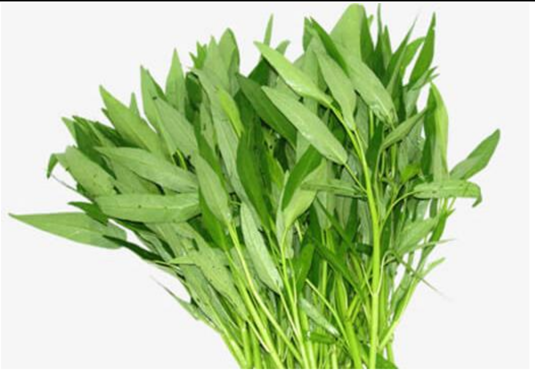 | 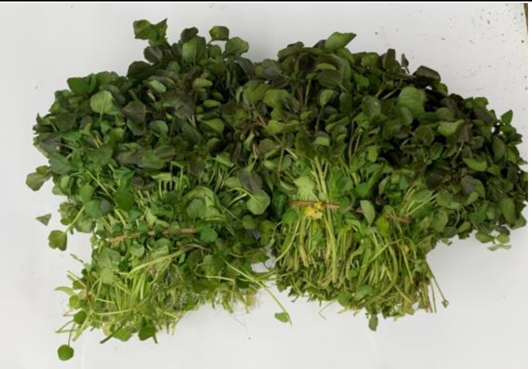 | 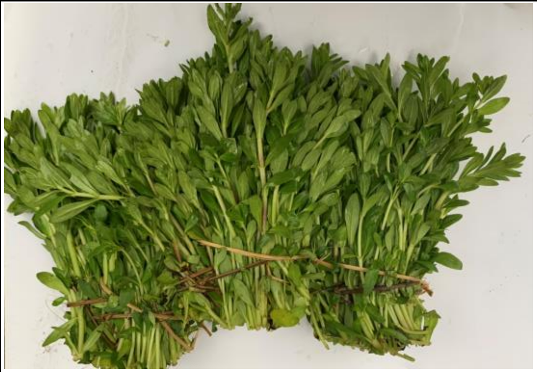 |
| Water spinach, water morning glory | Water cress | Cilantro |

### Water plants

| 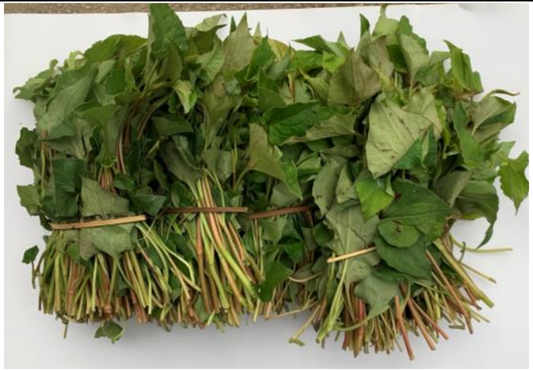 | 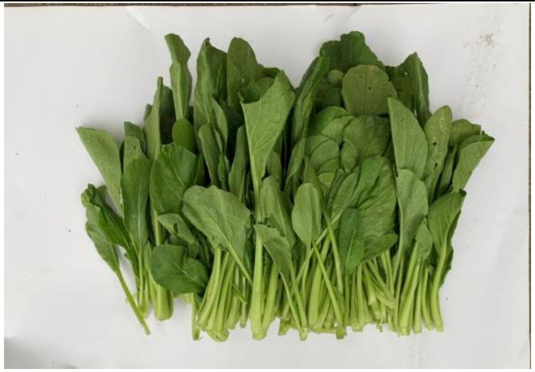 | 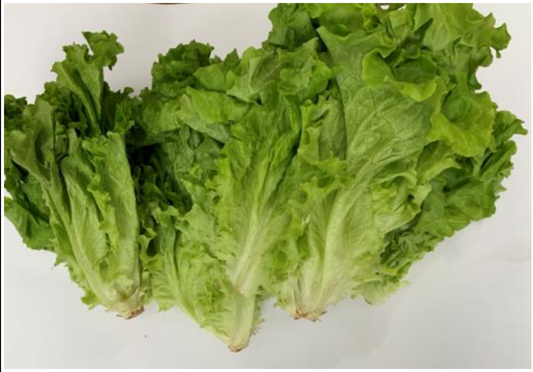 |
|---|---|---|
| Fish mint, lettuce mint | Sweet cabbage | Salad |

### Water plants

| | |
|---|---|
| 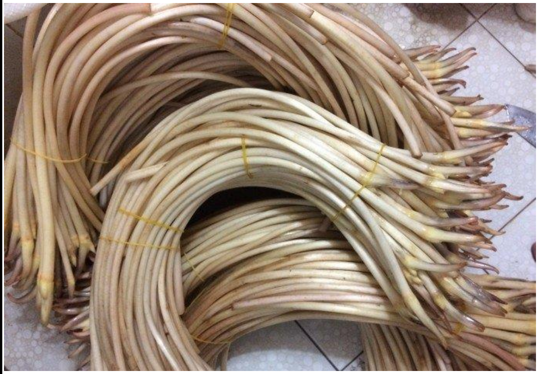 | 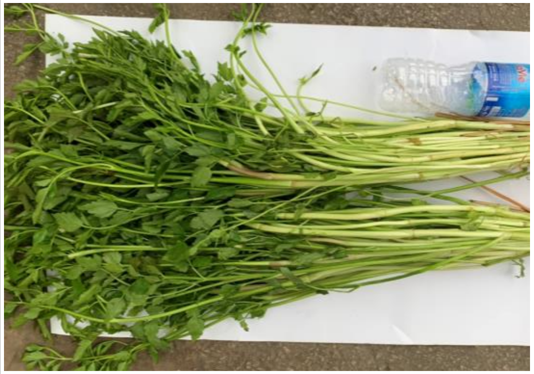 |
| Lotus | Celery |
